# Cognitive assessment methods and outcomes following shunt surgery in idiopathic normal pressure hydrocephalus (iNPH): a systematic review and meta-analysis

**DOI:** 10.64898/2026.01.25.26344784

**Authors:** Lisa M. Healy, Jeffrey Tooze, David Quist, Priya Varma, Christopher Carswell, Rocío Fernández-Méndez, John D. Pickard, Peter Smielewski, Alexis J. Joannides

## Abstract

**INTRODUCTION:** Core cognitive deficits in iNPH include slowed information processing, psychomotor slowing and executive dysfunction. However, the cognitive benefits of iNPH treatment with shunt surgery are not well understood. This review synthesised evidence on cognitive assessment methods and outcomes following shunt surgery in iNPH.

**METHODS:** PubMed, Scopus, PsycINFO and Web of Science were searched for peer-reviewed studies including adults with iNPH who underwent shunt surgery and had within-subject cognitive evaluations pre- and post-operatively. Key data were extracted and study quality was assessed. Random-effects meta-analyses were performed on pooled baseline and post-shunt difference scores for frequently reported cognitive tests with comparable data.

**RESULTS:** Of 1,876 records, 195 met the inclusion criteria, comprising 11,445 patients. Cognitive evaluation methods ranged from subjective reports and NPH grading scales to brief screening tools and comprehensive test batteries. Over 193 distinct tests were reported and 54.4% of studies did not formally assess any core iNPH cognitive deficits. Post-shunt improvement rates, follow-up times and criteria for defining improvement varied widely. Eighty-five studies contributed data to meta-analyses of ten outcomes. Pooled estimates indicated post-shunt cognitive improvement, with Trail Making Test-A, Grooved Pegboard-Dominant and Trail Making Test-B showing changes exceeding thresholds for clinically significant improvement.

**CONCLUSIONS:** Cognitive assessment in iNPH is highly heterogeneous and frequently omits core domains, limiting detection of treatment effects. When domain-relevant cognitive measures are used, shunt surgery is associated with statistically and clinically significant cognitive improvement. These findings highlight the need for standardised iNPH-specific cognitive evaluation tools with validated criteria for detecting clinically meaningful change and have direct implications for clinical assessment, interpretation of shunt response and the selection of cognitive endpoints in future interventional studies.

**Summary Box:** *What is already known on this topic:* Cognitive outcomes after shunt surgery for idiopathic normal pressure hydrocephalus (iNPH) have been inconsistently reported, with cognitive improvement reported less reliably than gait outcomes, in the context of highly variable assessment practices across centres.

*What this study adds:* This systematic review of 195 studies (11,445 patients) shows substantial heterogeneity in iNPH cognitive assessment and demonstrates that when tests sensitive to frontal–subcortical dysfunction are used, shunt surgery is associated with statistically and clinically meaningful cognitive improvement. Widely used dementia screening tools, including the MMSE and MoCA, show changes largely within expected practice-effect ranges and do not adequately capture core iNPH cognitive deficits.

*How this study might affect research, practice or policy:* These findings demonstrate the need to standardise cognitive assessment in iNPH using appropriate iNPH-specific tools with validated metrics for determining clinically meaningful improvement. This will enable robust trial endpoints and accurate evaluation of cognitive benefits of shunting in routine clinical practice.

## Introduction

Idiopathic normal pressure hydrocephalus (iNPH) primarily affects older adults, and is characterised by gait and balance disturbance, cognitive impairment and urinary incontinence in the context of ventriculomegaly (1,2). The pattern of cognitive impairment typically reflects frontal-subcortical dysfunction, with core deficits in information processing speed, psychomotor speed, executive functioning and apathy, which can impact daily functioning and quality of life (3–6).

iNPH is often underdiagnosed and misdiagnosed due to its overlapping symptoms with neurodegenerative conditions, and because gait, cognitive and bladder symptoms are common in older adults and may be mistaken for normal ageing (7–9). Surgical CSF diversion by shunt insertion can alleviate iNPH symptoms (10,11). Whilst gait changes have been widely documented, the effect of shunt surgery on cognitive symptoms is not well understood (12–14), and cognitive impairment has been cited as the least likely symptom to improve post-surgery (15). This uncertainty may reflect the high prevalence of comorbid neurodegenerative and cerebrovascular pathologies in iNPH patients, which can independently contribute to cognitive decline and attenuate measurable cognitive gains post-shunt (16–18). Accurate cognitive assessment may therefore be important for diagnostic accuracy and for detecting treatment response (13).

Currently, there is no internationally standardised approach to cognitive assessment in iNPH for clinical or research purposes, leading to variability in the tests administered across centres (19,20). On this background, the purpose of the current review was to provide a comprehensive and up-to-date synthesis of the literature on cognitive outcomes in iNPH. Specifically, the aims were to understand (i) how cognition is assessed pre-post shunt, (ii) the cognitive outcomes following shunt surgery and (iii) the methods for defining post-shunt cognitive improvement. Meta-analyses were conducted on the most frequently used tests with extractable cognitive data.

## Methods

### Search strategy

Electronic databases PubMed, Scopus, PsycINFO and Web of Science were searched from inception to January 2021, with the search subsequently updated to 25 September 2025. The following terms were used, adapted for each database as appropriate: (iNPH OR idiopathic normal pressure hydrocephalus OR NPH OR normal pressure hydrocephalus) AND (shunt* OR CSF diversion OR surgery) AND (cognition OR cognitive OR neuropsychological assessment OR neuropsychological outcome OR shunt outcome). Reference lists of the included studies were manually screened for additional eligible studies. The protocol was registered with PROSPERO (ID CRD42021296112). The review followed the Preferred Reporting Items for Systematic Reviews and Meta-Analyses (PRISMA) guidelines (21).

### Study selection

Two reviewers [LH, DQ] independently screened abstracts to identify articles to retrieve in full. Discrepancies were resolved by discussion involving a third reviewer [PV]. The inclusion criteria were: (1) peer-reviewed studies in English, (2) adults who underwent shunt surgery for suspected iNPH, (3) within-subject evaluations of cognition before and after shunt surgery, (4) specified cognitive evaluation method and (5) extractable cognitive outcomes post-surgery. Studies including multiple diagnoses were required to have independently extractable iNPH data. Exclusion criteria included: (1) case studies, (2) patients with secondary NPH or cases where the exact nature of hydrocephalus was unclear, (3) composite improvement scores in the iNPH symptom triad, with no extractable cognitive data and (4) conference proceedings.

### Data extraction

Data extraction was completed and validated by two reviewers [LH, JT]. Key descriptive data was extracted: first author, publication year, country, study design, follow-up timeframe, cognitive evaluation method, improvement criteria, sample size and sample characteristics (mean/median age, sex and symptom duration). For RCTs and designs involving multiple conditions or healthy controls, only pre-post shunt cognitive data from iNPH participants was extracted. Demographic and cognitive outcome data were not duplicated when the same patient cohort was reported across multiple publications.

### Quality assessment

Methodological quality was assessed using the Methodological Index for Non-Randomised Studies (MINORS) (22). Studies were evaluated across eight domains: study aim, selection bias (consecutive patients), prospective design, appropriate data collection methods, unbiased assessment (blinding), adequacy of follow-up, attrition and prospective sample size calculation, each rated as weak (0), moderate (1), or strong (2). In line with the focus of the present review, domains relating to study aims and outcome assessment were interpreted with reference to the assessment of cognitive outcomes (Supplementary Table 1).

### Meta analyses

Meta-analyses were conducted on the pooled baseline and difference scores for the following tests: Digit Span Backwards (DSB), Digit Span Forwards (DSF), Frontal Assessment Battery (FAB), Grooved Pegboard Test-Dominant hand (GPB-D), Mini-Mental State Examination (MMSE), Montreal Cognitive Assessment (MoCA), Rey Auditory Verbal Learning Test (RAVLT), Trail Making Test-A (TMT-A) and Trail Making Test-B (TMT-B). These tests were selected based on having at least five studies with extractable mean (SD) or median (IQR, range) data from comparable test versions. Separate outcome data were available for the RAVLT Learning and Delayed Recall subtests, which were treated as distinct cognitive outcome measures for meta-analysis purposes. Insufficient comparable data were available for the Stroop and Phonemic Fluency tests due to heterogeneity in test versions, languages, scoring procedures or incomplete reporting of relevant summary statistics (23). For studies with multiple follow-ups, data from the last available post-shunt assessment were used. Data from multiple publications were not duplicated. Analyses were performed using IBM SPSS v30.

Where reported, medians (IQR, range) were converted to means (SD) using the Wan method (24), with Box-Cox transformations applied to skewed data (25). For case-series studies reporting individual pre- and post-shunt scores, group means and SDs were calculated manually from the raw data provided.

To account for variability across studies, random-effects meta-analyses were performed using the Restricted Maximum Likelihood (REML) method (26). Pooled difference scores were used as outcomes to account for sample sizes and variances. Effect sizes were expressed as unstandardised mean differences. Heterogeneity was assessed using the I² statistic (27). Publication bias was examined using funnel plots and Egger’s regression test (28). Trim-and-fill was applied where indicated, to estimate the impact of potentially missing studies (29). Results are presented as pooled difference scores with 95% confidence intervals.

### Clinical significance

Clinically significant improvement was defined a priori using two approaches: (i) age-matched, distribution-based thresholds derived from pooled normative standard deviations (0.3 SD = small, 0.5 SD = moderate, 1.0 SD = large clinical change), and (ii) age-corrected z-score change relative to normative data (≥1 z-score). These criteria allow statistically significant pooled effects to be interpreted against benchmarks of clinically meaningful cognitive improvement (30–34).

### Sensitivity analyses

Sensitivity analyses were conducted to assess the robustness of pooled estimates and the influence of outlier or high-variance studies. Stepwise exclusions were performed for each cognitive outcome, prioritising studies with extreme mean differences, large standard errors, or visually identified influence in forest or funnel plots. Outlier studies were also identified based on conceptual and psychometric criteria, such as marked directional inconsistency with other outcomes within the same study, or extreme variance shifts. After each exclusion, random-effects models were re-estimated as described above. Sensitivity analyses were considered confirmatory when the direction and significance of pooled effects were preserved (23,35).

## Results

### Search identification

After removing duplicates, 1,876 unique records were identified from the systematic search (Figure 1). Titles and abstracts were screened and 1,386 records were excluded. One article was not accessible for full text review or retrievable from the corresponding author. Full texts of the remaining 489 studies were assessed for eligibility. An additional 31 studies were identified from backwards citation searching. In total, 195 studies met the inclusion criteria and were included in the final review. A full reference list is provided in the Supplementary Material. Study characteristics and participant demographics are summarised in Table 1.

**Figure 1.**
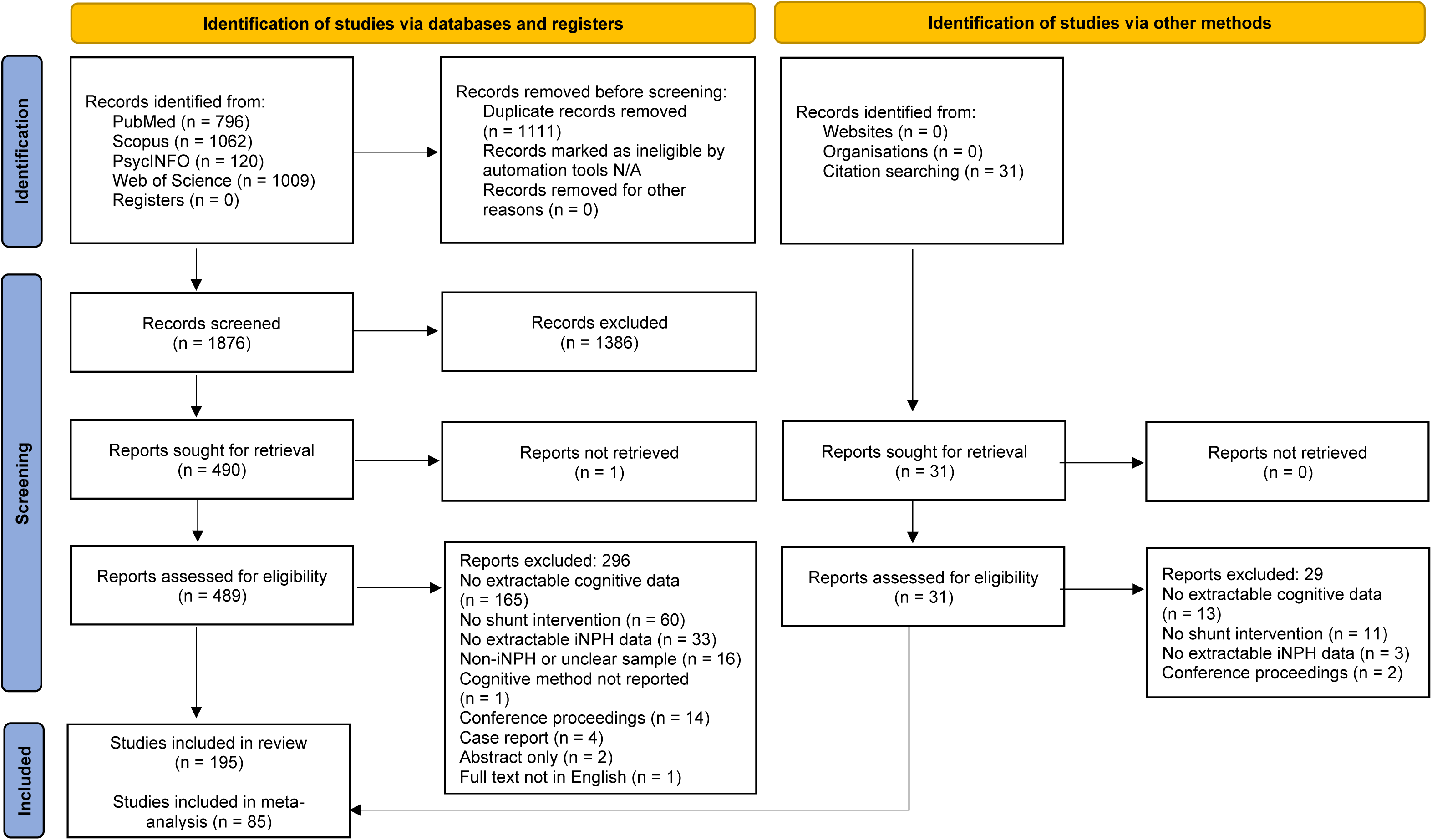
PRISMA diagram.

**Table 1.** Study and participant characteristics.

| Characteristic | Summary |
| --- | --- |
| Total studies (n) | 195 |
| Study design, n (%) |  |
| Prospective | 118 (60.5%) |
| Retrospective | 77 (39.5%) |
| Participants (n) | 11,445 |
| Sex, n (%) |  |
| Male | 6,459 (56.4%) |
| Female | 4,555 (39.8%) |
| Unspecified | 431 (3.8%) |
| Median age (years) | 70–77 |
| Symptom duration at baseline |  |
| Median | 24 months |
| Range | 1–72 months |
| Countries represented (n = 26) | Argentina, Australia, Austria, Belgium, Brazil, Canada, China, Czechia, Denmark, Finland, Germany, Greece, Hungary, Italy, Japan, Netherlands, Norway, Portugal, Republic of Korea, Spain, Sweden, Switzerland, Taiwan, Turkey, UK, USA |

**Note.** Median age and symptom duration were available for subsets of studies.

### Quality assessment (MINORS)

Application of the MINORS criteria demonstrated heterogeneous methodological quality across domains (Figure 2; Supplementary Table 1). Weak ratings were most frequently observed for unbiased outcome assessment (blinding) and prospective sample size calculation. Prospective design, consecutive patient inclusion and adequacy of follow-up were more commonly rated as moderate to strong.

**Figure 2.**
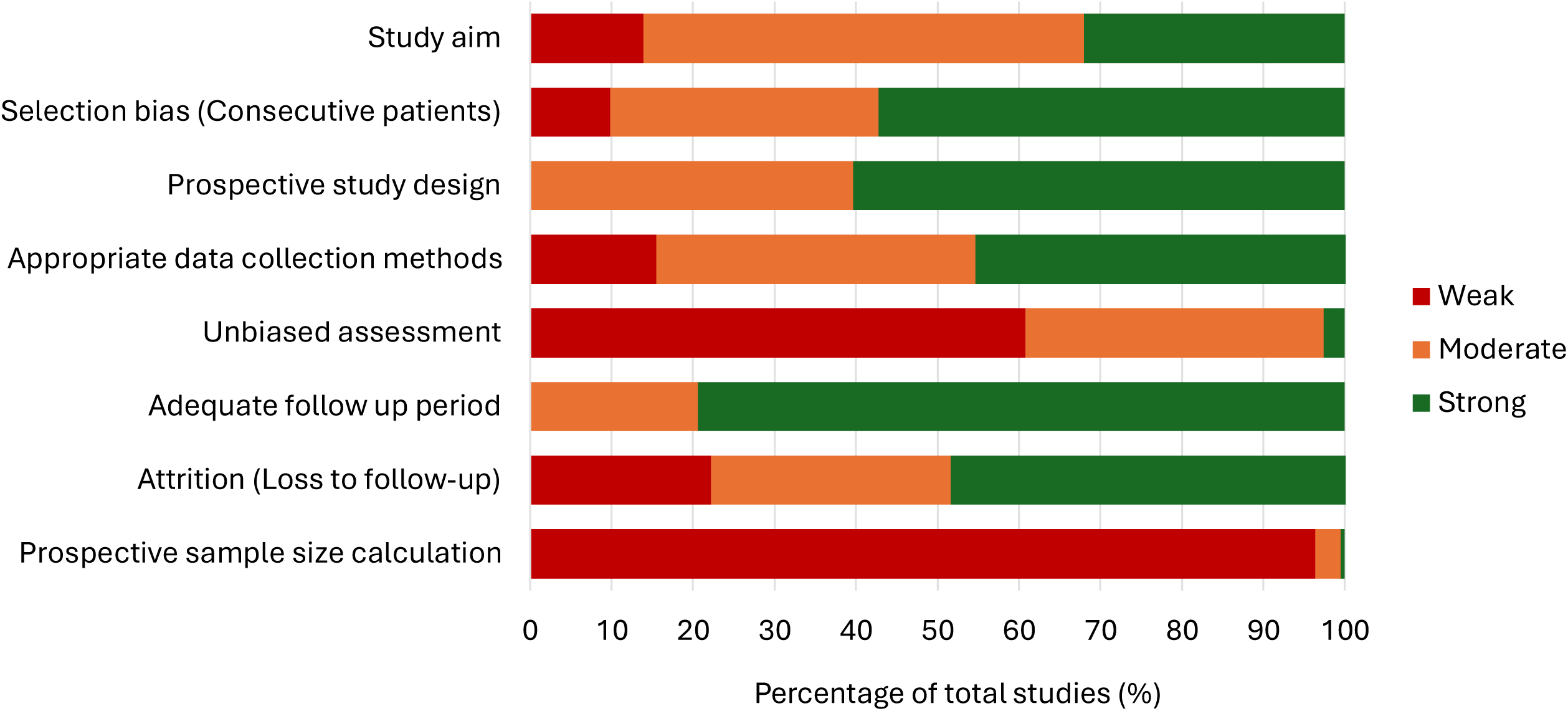
Distribution of MINORS quality assessment ratings across methodological domains in the 195 included studies.

### Cognitive evaluation pre-post shunt

#### Timing of testing

Of the 195 included studies, 137 (70.3%) reported cognitive data from a single post-operative follow-up, 29 (14.9%) reported two follow-ups, 19 (9.7%) reported three or more follow-ups (up to a maximum of six) and 10 (5.1%) did not specify the follow-up time used. Modal follow-up times were 3 and 12 months (Range 2 days to 8-10 years post-shunt). The distribution of follow-up intervals is presented in Figure 3.

**Figure 3.**
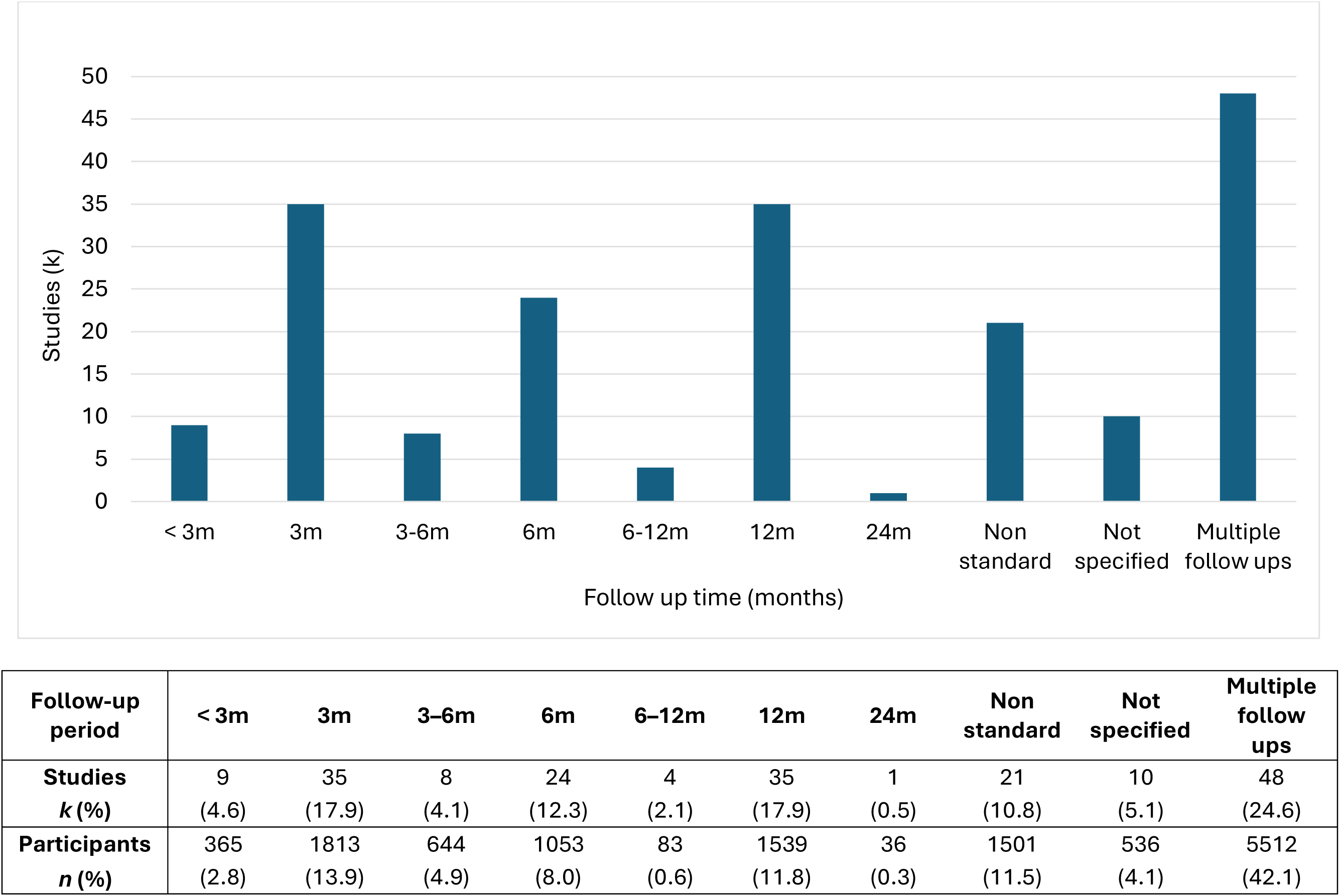
Distribution of follow-up times in 195 studies with post-shunt cognitive data.

#### Cognitive evaluation methods

Subjective ratings by patients, clinicians and/or carers were reported in 89 studies (45.6%). Subjective ratings were used as a standalone in 30 studies (15.4%) and in combination with cognitive tests in 59 (30.3%) studies. Standardised symptom measures or structured interviews were primarily used with carers (13/20 studies).

61 studies (31.3%) had extractable cognitive data from NPH grading scales, including various iterations of the Japanese NPH Grading Scale–Cognition (JNPHGS-Cognition; 39/195, 20.0%), Hellstrom iNPH Grading Scale-Neuropsychology (12/195, 6.2%) and the Sahuquillo NPH Scale-Cognitive Function (10/195, 5.1%). Respectively, the NPH grading scales were based on subjective clinician ratings, a composite score generated from cognitive tests, or a combination of subjective and objective test data. A further seven studies used author-specific grading scales.

165 studies (84.6%) used at least one cognitive testing method. Brief non-specific dementia screening tools (e.g. MMSE, MoCA) were used as standalone cognitive tests in 63 studies (32.3%). 102 studies (52.3%) used cognitive test batteries containing two or more tests. Test batteries were composed of a median of five tests (IQR 3-10, range 2-33). Two studies did not specify the final number of tests used.

Table 1 lists the most frequently reported cognitive tests, with extractable cognitive outcome data. At least 193 distinct cognitive tests were identified across the included studies, 127 of which appeared in one study only. These tests assessed a broad range of cognitive functions spanning attention and working memory, executive function, language and verbal fluency, memory and orientation, information processing speed, psychomotor speed, reaction time, social cognition and visuospatial function. The reported cognitive tests are listed in full in Supplementary Table 2.

**Table 1.** Most frequently used cognitive tests with extractable data.

| Test | Main cognitive domains | Studies |  | Participants |  |
| --- | --- | --- | --- | --- | --- |
|  |  | *k* | (%) | *n* | (%) |
| MMSE | Dementia screen | 128 | (65.6) | 8227 | (71.9) |
| TMT-A | Psychomotor / processing speed | 38 | (19.5) | 1342 | (11.7) |
| FAB | Executive function screen | 35 | (17.9) | 969 | (8.5) |
| RAVLT | Memory | 32 | (16.4) | 1408 | (12.3) |
| Stroop Interference | Executive function | 32 | (16.4) | 1334 | (11.7) |
| Grooved Pegboard Test | Psychomotor speed | 27 | (13.8) | 1170 | (10.2) |
| Digit Span Backward | Attention / working memory | 26 | (13.3) | 993 | (8.7) |
| Digit Span Forward | Attention | 26 | (13.3) | 993 | (8.7) |
| TMT-B | Executive function | 25 | (12.8) | 971 | (8.5) |
| Semantic Fluency | Verbal fluency / language | 22 | (11.3) | 804 | (7.0) |
| Stroop Colour Naming | Processing speed | 22 | (11.3) | 1293 | (11.3) |
| Phonemic Fluency | Verbal fluency / executive function | 20 | (10.3) | 585 | (5.1) |

**Note.** Percentages refer to the proportion of total studies ( $k = 195$ ) and participants ( $n = 11,445$ ). FAB = Frontal Assessment Battery, MMSE = Mini-Mental State Examination, TMT = Trail Making Test, RAVLT = Rey Auditory Verbal Learning Test. Digit Span, Phonemic Fluency, RAVLT, Semantic Fluency, Stroop, TMT-A and TMT-B include all test versions and languages.

More than half of the shortlisted studies (106/195, 54.4%), did not formally test any of the core cognitive deficits observed in iNPH. 58 studies (29.7%) reported test batteries including both a measure of executive function and processing speed or psychomotor speed, which are core cognitive deficits observed in iNPH. Seven studies reported a measure of reaction time, a related indicator of the cognitive slowing characteristic of iNPH. The most common cognitive evaluation tool was the MMSE dementia screen, with extractable cognitive data in 128 studies (65.6%). Regarding test modality, most studies used conventional paper and pencil methods to administer cognitive tests. Nine studies (4.6%) reported a computerised assessment method.

### Definition of post-shunt cognitive improvement

Post-shunt cognitive improvement (PCI) was defined using heterogenous criteria across studies. PCI was operationalised by improvement rate in 56 studies (28.7%), statistically significant sample-level improvement in 92 studies (47.2%) and a combination of both approaches in 47 studies (24.1%), based on at least one post-operative follow-up. Of the 103 (52.8%) studies that defined PCI using an improvement rate approach, 42 exclusively used subjective ratings, 52 used objective testing and nine incorporated both methods.

Subjective ratings of PCI were reported either as dichotomous outcomes (improvement, no improvement; n = 43) or stratified using graded improvement scales (range 3-4 points; n = 8) rated by clinicians/researchers. For the JNPHGS-Cognition, the most common improvement threshold was ≥1 point.

Considerable variability was observed in the operational definitions of PCI among studies reporting objective test data. Most studies specified diagnostic rules applied to individual cognitive tests or test batteries. The most common criterion was improvement by a specified number of points or reduction in completion time (36/58 studies). 17 studies applied two or more diagnostic rules to define PCI. A validated improvement metric was reported in eight studies (z-score, t-score or SD change). Table 2 outlines the diagnostic rules applied.

**Table 2.** Diagnostic rules used to define post-shunt cognitive improvement based on individual cognitive tests or test batteries in 58 studies.

| **Individual cognitive tests** | **Studies**<br>**(n)** |
| --- | --- |
| Test improved by: |  |
| Percentage (Range 5-25%) | 5 |
| Specified points or completion time | 36 |
| $\geq 1$ SD | 2 |
| $\geq 5$ T-Score points | 1 |
| Test improved to a specific points threshold | 1 |
| Criteria NS | 10 |
| **Batteries of cognitive tests** | **Studies**<br>**(n)** |
| $\geq 1$ SD improvement on $\geq 1/12$ tests | 1 |
| $\geq 1$ SD improvement on $\geq 50\%$ tests | 3 |
| $\geq 25\%$ improvement on $\geq 50\%$ of tests | 1 |
| $\geq 33.3\%$ of tests in battery improved (Score NS) | 1 |
| $\geq 50\%$ of tests in battery improved (Score NS) | 2 |
| Composite score improved (Criteria NS) | 6 |
| Composite score improved by specified points | 2 |
| Composite score improved by $\geq 1$ z-score | 1 |
| Executive function tests improved (Criteria NS) | 1 |
| Criteria NS | 4 |

**Note.** 17 studies applied two or more diagnostic rules; SD = standard deviation; NS = not specified

### Post-shunt cognitive outcomes

Of the 139 studies defining PCI by statistical significance, 110 (79.1%) reported improvement in at least one cognitive parameter. Among studies reporting an improvement rate, the pooled PCI rate was 56.6% for subjective reports (range 12.5-87.5%) and 48.4% for cognitive testing (range 11.8-100%). Five studies also reported PCI rate based on combined subjective and objective data, yielding a pooled rate of 64.2% (range 39.5-85.5%). Pooled PCI rate at each follow-up period is presented in Figure 3.

### Meta analyses of post-shunt cognitive change

Eighty-five studies met the inclusion criteria for the meta-analyses (Table 3). Extractable cognitive outcome data from ≥5 studies were available for DSB, DSF, FAB, GPB-D, MMSE, MoCA, RAVLT Learning, RAVLT Delayed Recall, TMT-A and TMT-B. Heterogeneity ranged from low to substantial (I² = 9.4–64.3%), with no outcome reaching the threshold for considerable heterogeneity. Egger’s test indicated publication bias for the MMSE (*p* = .029), with no evidence of bias for other measures. Trim-and-fill adjustment resulted in modest attenuation of pooled effects, which remained statistically significant.

**Table 3.** Meta analyses results for 85 studies.

| Cognitive outcome measure | Studies (k) | Estimated average pre-shunt score | Estimated average difference score | 95 % CI | p | Cochran's Q (df; p) | I<sup>2</sup> (%) | Between-study variance ($\tau^2$) | Egger's test (p) | Clinically significant |
| --- | --- | --- | --- | --- | --- | --- | --- | --- | --- | --- |
| **Executive function, attention and working memory** |  |  |  |  |  |  |  |  |  |  |
| DSB | 10 | 2.95 digits | 0.33 digits | [0.10, 0.55] | 0.005 | 26.24 (9; 0.002) | 64.3 | 0.07 | 0.781 | No |
| DSF | 10 | 4.69 digits | 0.10 digits | [-0.03, 0.24] | 0.129 | 6.34 (9; 0.705) | 9.4 | 0.01 | 0.944 | No |
| FAB | 24 | 11.18 points | 1.56 points | [1.04, 2.08] | <.001 | 69.04 (23; <.001) | 60.0 | 0.92 | 0.217 | No |
| TMT-B | 13 | 234.44 seconds | -35.18 seconds | [-59.91, -10.46] | 0.005 | 25 (12; 0.015) | 47.9 | 880.19 | 0.990 | Yes |
| **Processing speed / psychomotor speed** |  |  |  |  |  |  |  |  |  |  |
| GPB-D | 5 | 155.25 seconds | -31.54 seconds | [-43.47, -19.61] | <.001 | 3.74 (4; 0.442) | 10.7 | 20.46 | 0.272 | Yes |
| TMT-A | 21 | 114.74 seconds | -22.73 seconds | [-31.35, -14.11] | <.001 | 22.05 (20; 0.338) | 16.6 | 60.41 | 0.252 | Yes |
| **Learning and memory** |  |  |  |  |  |  |  |  |  |  |
| RAVLT Learning | 12 | 23.70 words | 5.58 words | [3.77, 7.39] | <.001 | 18.65 (11; 0.068) | 41.3 | 3.50 | 0.122 | No |
| RAVLT Delayed | 11 | 2.75 words | 1.37 words | [0.86, 1.88] | <.001 | 16.62 (10; 0.083) | 42.9 | 0.26 | 0.149 | No |
| **Global cognitive function** |  |  |  |  |  |  |  |  |  |  |
| MoCA | 7 | 19.06 points | 1.75 points | [0.79, 2.71] | <.001 | 9.41 (6; 0.152) | 22.4 | 0.37 | 0.387 | No |
| MMSE | 71 | 22.94 points | 1.57 points | [1.23, 1.91] | <.001 | 149.95 (70; <.001) | 61.0 | 0.95 | 0.029 | No |
| MMSE (imputed) | 88 | - | 1.07 points | [0.67, 1.47] | <.001 | - | - | - | - | - |

All outcome measures showed pooled difference scores in the direction of cognitive improvement following shunt surgery. Statistical significance was therefore interpreted alongside predefined thresholds for clinically meaningful change. The magnitude of pooled effects varied across tests, with DSF (*p* = .129) the only outcome that did not reach statistical significance. Among the nine significant outcomes, mean difference scores for six measures (DSB, FAB, MMSE, MoCA, RAVLT Learning and RAVLT Delayed Recall) fell within ranges typically attributed to practice effects, test–retest variability, or measurement error, rather than indicating genuine post-shunt cognitive change (36,37).

Three measures, TMT-A, GPB-D and TMT-B (for which faster completion time indicates better performance), demonstrated statistically and clinically significant improvements that exceeded all prespecified thresholds for clinically meaningful change (30–34). Figure 4 presents the forest plots for TMT-A, GPB-D and TMT-B. Remaining outcomes are displayed in Supplementary Figures 1-7.

**Figure 4.**
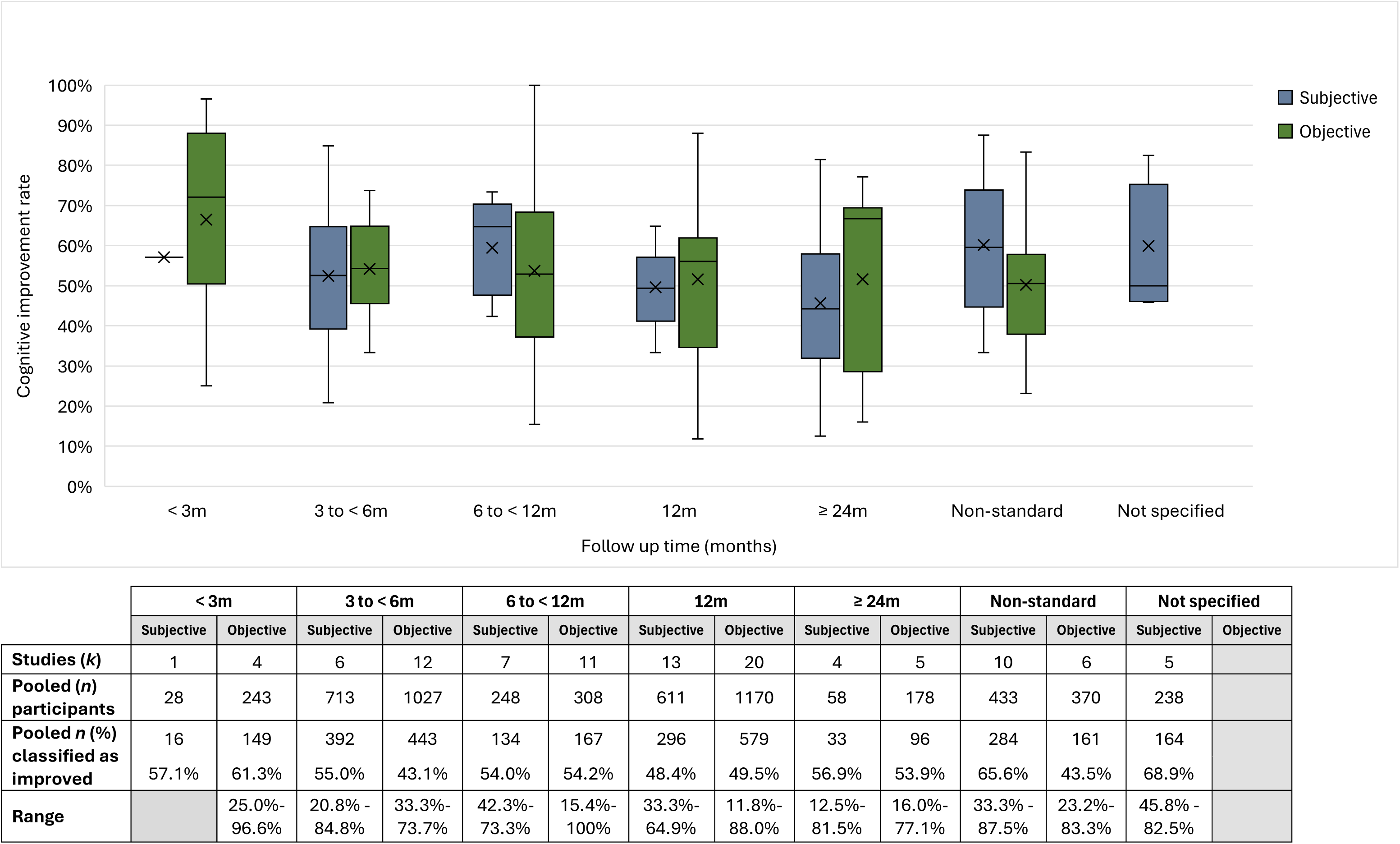
Pooled post-shunt cognitive improvement (PCI) rate in studies with extractable data. Box plots represent the mean, median and 25th and 75th percentiles for subjective and objective data at each follow up period. Whiskers represent the range.

Sensitivity analyses confirmed that pooled cognitive effects were robust across outcomes. Stepwise exclusion of influential or outlier studies and trim-and-fill analyses did not materially alter the direction or significance of any pooled estimates (Supplementary Table 3).

**Figure 5.**
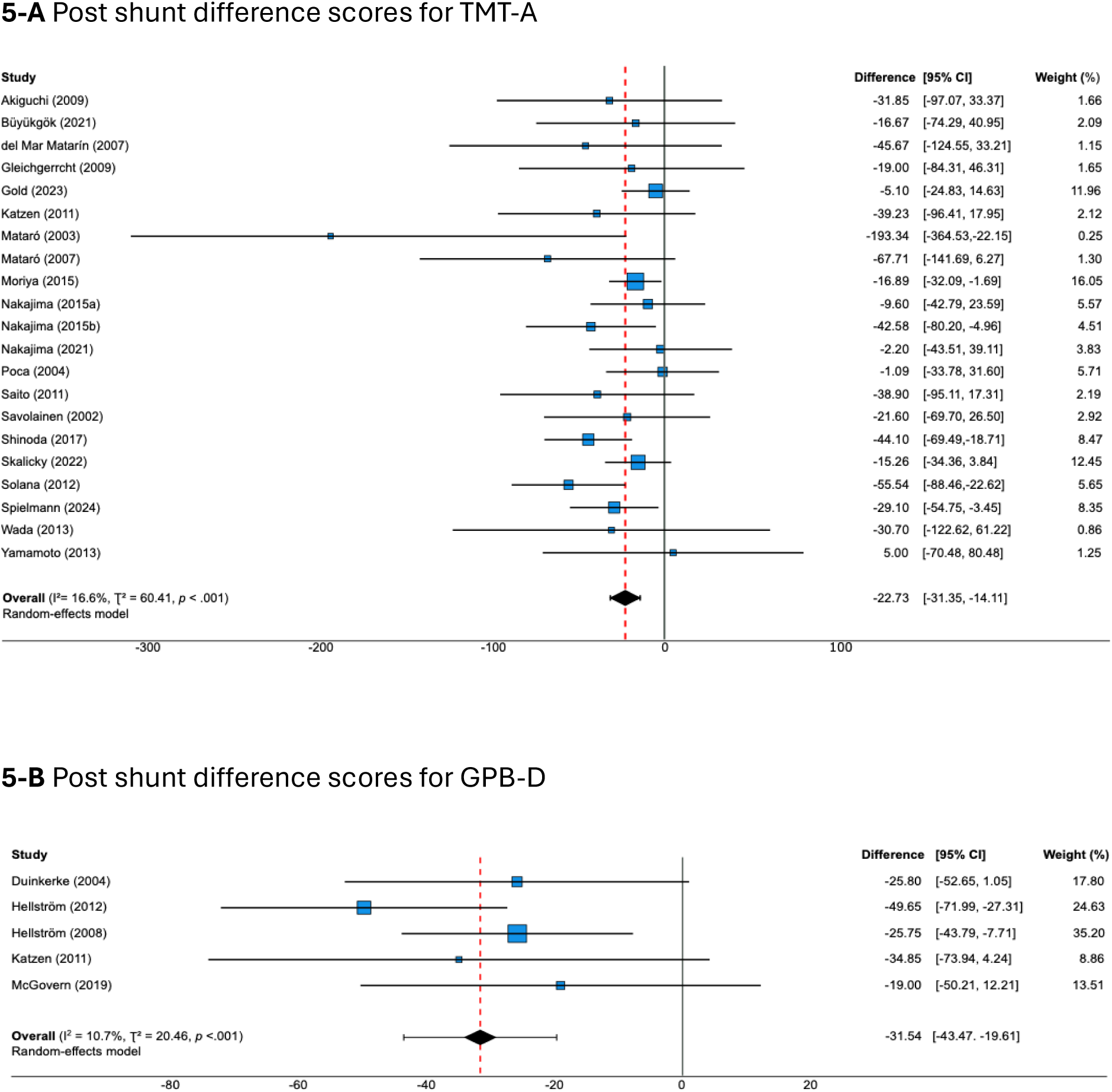
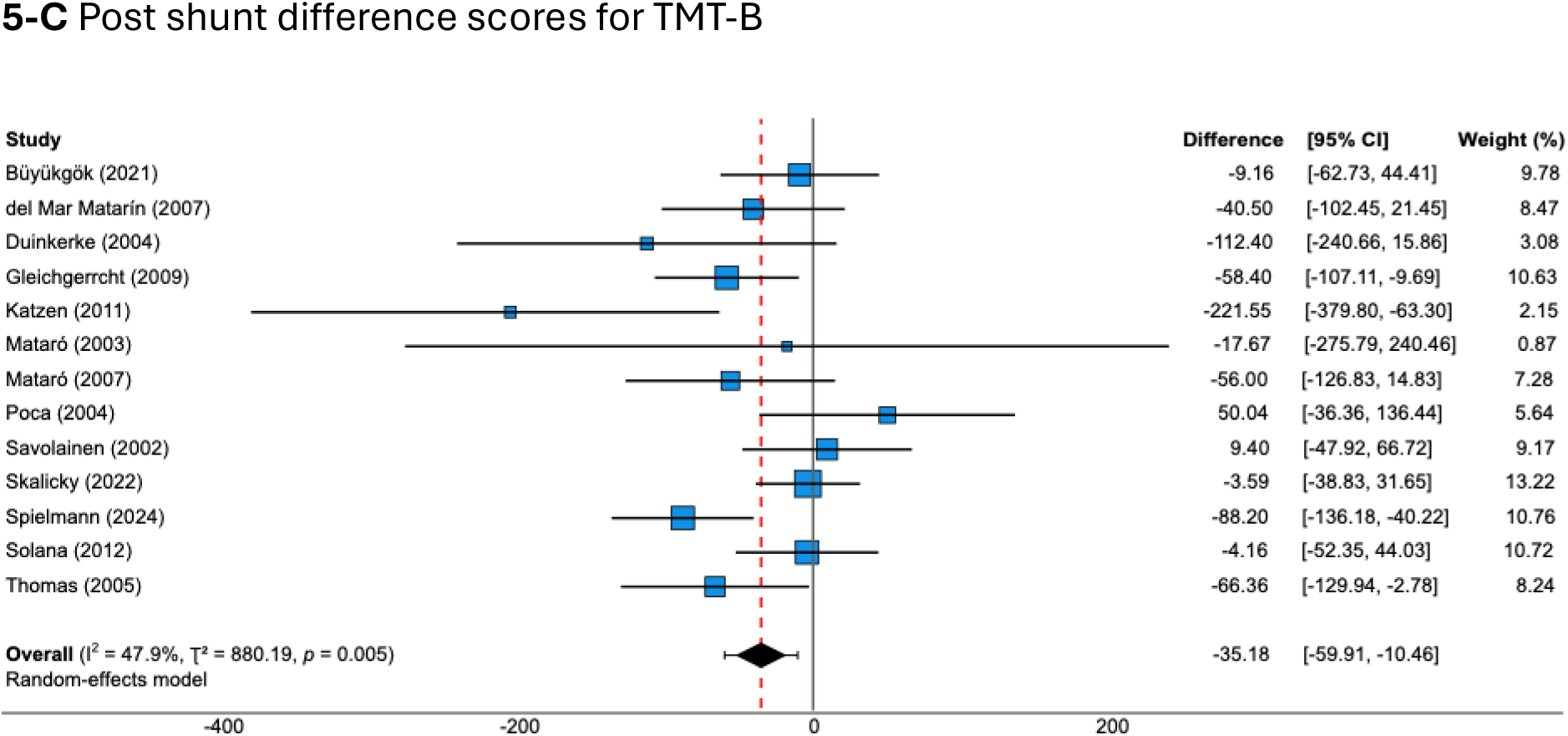
Forest plots of post-shunt difference scores for TMT-A, GPB-D and TMT-B. **Note.** Pooled mean difference scores were calculated using a random-effects model. Mean difference estimates for individual studies are presented with corresponding 95% confidence intervals and percentage weights. For each test, the unit of measurement is completion time in seconds. Negative mean differences indicate improved performance (faster completion).

## Discussion

This systematic review represents the largest pooled iNPH cohort evaluating post-shunt cognitive outcomes, assessment methods and definitions of cognitive improvement. Drawing on 195 studies, encompassing 11,445 patients from 26 countries, it combines subjective and objective assessment approaches to provide a comprehensive synthesis of cognitive change following shunt surgery. Critically, this review reveals that substantial methodological heterogeneity may have obscured true treatment effects. Importantly, when measures targeting core iNPH cognitive domains are employed, there is evidence of statistically and clinically significant cognitive improvement following shunt surgery.

### Heterogeneity in cognitive assessment and improvement criteria

Considerable heterogeneity was observed in both cognitive assessment methods and definitions of post-shunt cognitive improvement. Cognitive evaluation varied widely in test selection and timing of post-operative reassessment, with most studies (70.3%) reporting a single cognitive follow-up and few including longitudinal data. This is an important consideration given the chronic and progressive nature of iNPH, in which cognitive outcomes may evolve over time (9).

Test battery composition was highly heterogeneous, with 127 of 193 reported tests used only once. More than half of studies failed to assess any core iNPH-related cognitive domains (processing speed, psychomotor speed and executive function), reflecting the absence of a standardised assessment approach. Commonly used dementia screening tools, such as the MMSE, were frequently used despite poor sensitivity to frontal–subcortical dysfunction and limited ability to detect change due to ceiling effects (38). Furthermore, sensitive measures were embedded within global composite scores in several studies, potentially masking domain-specific improvement. This aligns with a review of four RCTs, which also reported a lack of assessment uniformity and noted that commonly used tools lack sensitivity to NPH-relevant cognitive change (12).

Heterogeneity in how cognitive improvement was defined further compounded these assessment issues. Many studies relied solely on statistical significance as evidence of improvement, potentially overlooking clinically meaningful individual-level change. Few studies stratified analyses by baseline cognitive impairment, despite not all patients with iNPH presenting with cognitive deficits (2). This may have diluted observable post-shunt effects among those with bassline cognitive deficits and obscured clinically meaningful change.

Arbitrary thresholds were commonly applied to define improvement rate, with only eight studies using recognised approaches such as z-score or SD-based change, and none reporting use of the reliable change index (33,34). Subjective cognitive improvement was often poorly defined, with most studies relying on dichotomised outcomes and unstructured ratings, rendering them insensitive to subtle change and vulnerable to recall bias (39). Although the JNPHGS-Cognition offers a structured grading system, its categories are not specific to iNPH and may be most useful when interpreted alongside targeted cognitive testing (40).

Together, this methodological and definitional heterogeneity resulted in variability in reported improvement rates, including wide variation within objectively assessed outcomes. This suggests that test selection and improvement criteria are key contributors to outcome heterogeneity, which could potentially obscure true treatment response.

### Evidence for cognitive improvement with appropriate testing

Despite substantial methodological heterogeneity across studies, the meta-analysis demonstrates that shunt surgery is associated with clinically meaningful cognitive improvement when tests sensitive to iNPH-related deficits are used. Among the ten cognitive outcomes assessed, TMT-A, GPB-D and TMT-B showed statistically significant improvements that exceeded expected practice effects and all prespecified thresholds for clinically meaningful change (30–34). Sensitivity analyses confirmed the robustness of these findings.

TMT-A and GPB demonstrated low heterogeneity indicating consistent effects across studies. Although TMT-B showed moderate-substantial heterogeneity, this may partly reflect inconsistent application of the test’s 5-minute discontinuation rule (32), which patients with severe processing speed deficits are more likely to reach, potentially inflating between-study variability. Together, these results indicate that when validated measures targeting core iNPH cognitive deficits are used, shunt surgery may yield reliable and clinically meaningful cognitive improvement.

### Limitations

Potential limitations include variability in post-shunt follow-up timing and the reliance on single post-operative assessments, limiting insight into cognitive recovery trajectories. Methodological heterogeneity in cognitive evaluation tools and definitions of improvement may also reduce comparability and generalisability across studies.

Meta-analysis of several commonly used executive function (Phonemic Fluency, Stroop) and processing speed tests (SDMT, Digit Symbol/Coding) was not possible due to insufficient reporting of relevant summary statistics and heterogeneity in test versions or scoring methods. The GPB-D meta-analysis was limited by the small number of extractable datasets, although effect direction and magnitude were consistent across studies.

Study quality was variable, with frequent limitations relating to lack of prospective sample size calculation or blinded outcome assessment. In addition, restricting search inclusion to English-language publications may have introduced geographical bias. Despite these limitations, this review represents the most comprehensive synthesis to date of cognitive assessment methods and post-surgical outcomes in iNPH, identifying key methodological factors that have systematically influenced outcome reporting in this field.

### Clinical practice implications

Although cognitive impairment has been cited as the least responsive symptom of Hakim’s triad following shunt surgery (15), this review suggests that previously inconsistent findings may, at least in part, reflect methodological limitations rather than true treatment ineffectiveness. In particular, more than half of studies failed to assess domains most relevant to iNPH or relied on tests with limited sensitivity to change, which has clear implications for clinical assessment.

The findings suggest that incorporating cognitive tests sensitive to frontal–subcortical dysfunction may improve the evaluation of cognitive outcomes in iNPH. Brief measures of processing speed, psychomotor speed and executive function, such as TMT-A, GPB-D and TMT-B, were associated with statistically and clinically meaningful improvements in the meta-analyses, although the limited number of studies reporting extractable GPB-D data warrants cautious interpretation.

Commonly used screening tools (MMSE, MoCA, FAB) do not adequately assess core iNPH-related cognitive deficits and show changes largely within practice-effect ranges, limiting their utility for evaluating shunt response. This may partly explain the absence of cognitive improvement reported in a recent RCT using the MoCA as the primary outcome (14). Widely used dementia screening tools (MMSE, MoCA, ACE-III) may be better suited for differential diagnosis or identifying cognitive comorbidities in complex NPH presentation. Common iNPH comorbidities such as Alzheimer’s disease or vascular pathology can independently impair cognition and attenuate observable post-shunt improvement, making their detection essential for accurate interpretation of cognitive outcomes (16–18).

More broadly, these findings highlight the need for greater standardisation in cognitive assessment for iNPH. Consistent use of domain-relevant tests, alongside validated metrics for defining clinically meaningful change, has the potential to improve diagnostic accuracy, evaluation of shunt response and the long-term management of patients with iNPH (13).

## Conclusion

This systematic review demonstrates that when appropriate cognitive assessment tools are used, there is evidence of clinically meaningful post-shunt cognitive improvement in iNPH. Much of the inconsistency in prior literature reflects considerable methodological heterogeneity in cognitive assessment and improvement criteria, rather than treatment ineffectiveness. Many commonly used tests fail to capture the core cognitive deficits observed in iNPH, limiting their utility for assessing baseline impairment and post-shunt change.

These findings emphasise the need for standardised, iNPH-specific cognitive evaluation tools that are feasible for routine practice and incorporate validated metrics for detecting clinically relevant cognitive improvement. Future research should prioritise the development and validation of such tools, including measures of processing speed, psychomotor speed and executive function, with longitudinal follow-up to capture the trajectory of post-shunt cognitive recovery. Importantly, implementation of such standardised approaches has the potential to improve patient selection, enhance surgical outcomes and optimise long-term management for this under-recognised yet treatable cause of dementia.

## Data Availability

All data produced in the present work are contained in the manuscript

## Supplementary Materials

Cognitive assessment methods and outcomes following shunt surgery in idiopathic normal pressure hydrocephalus (iNPH): a systematic review and meta-analysis

**Supplementary Table 1.**
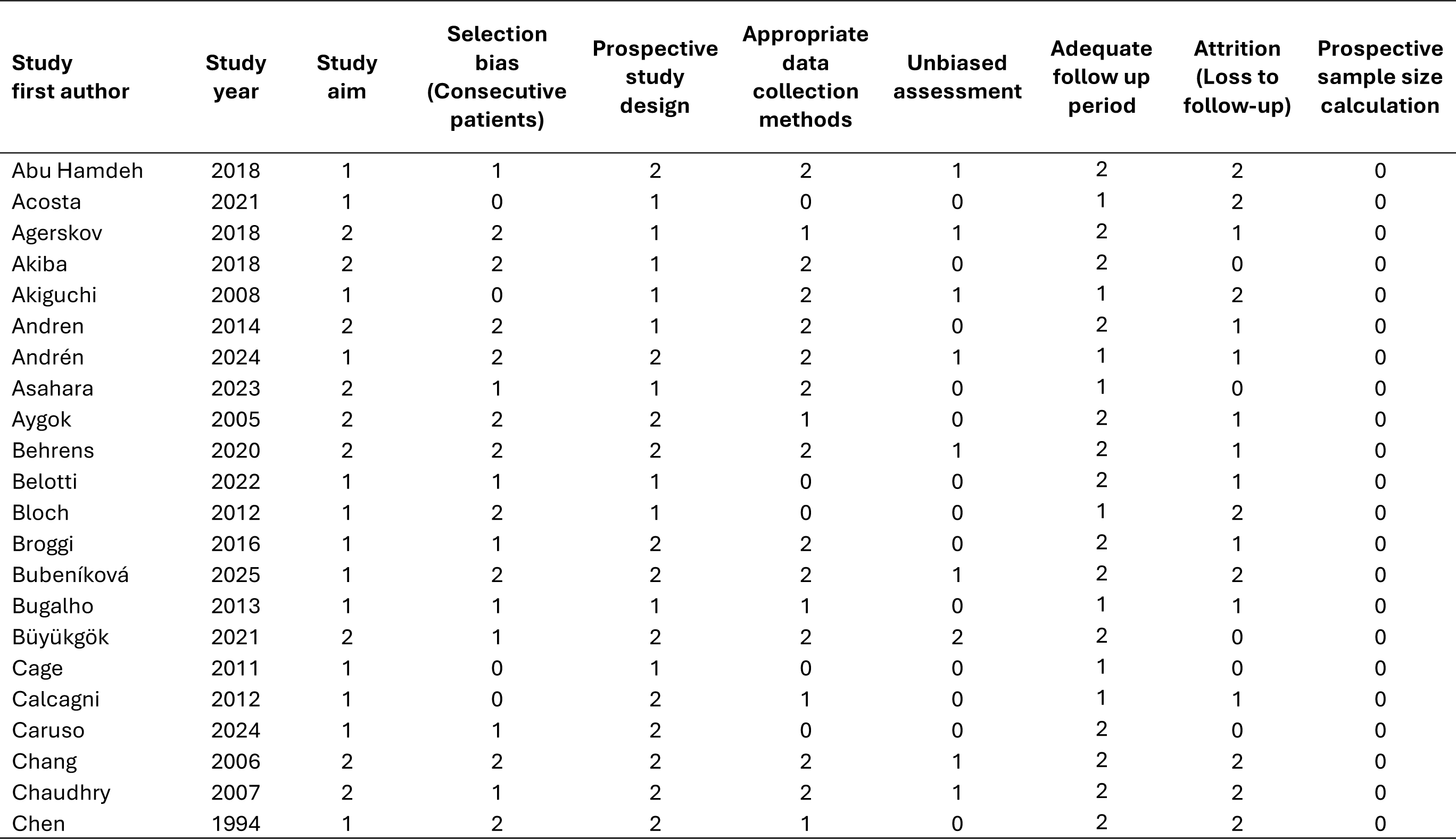
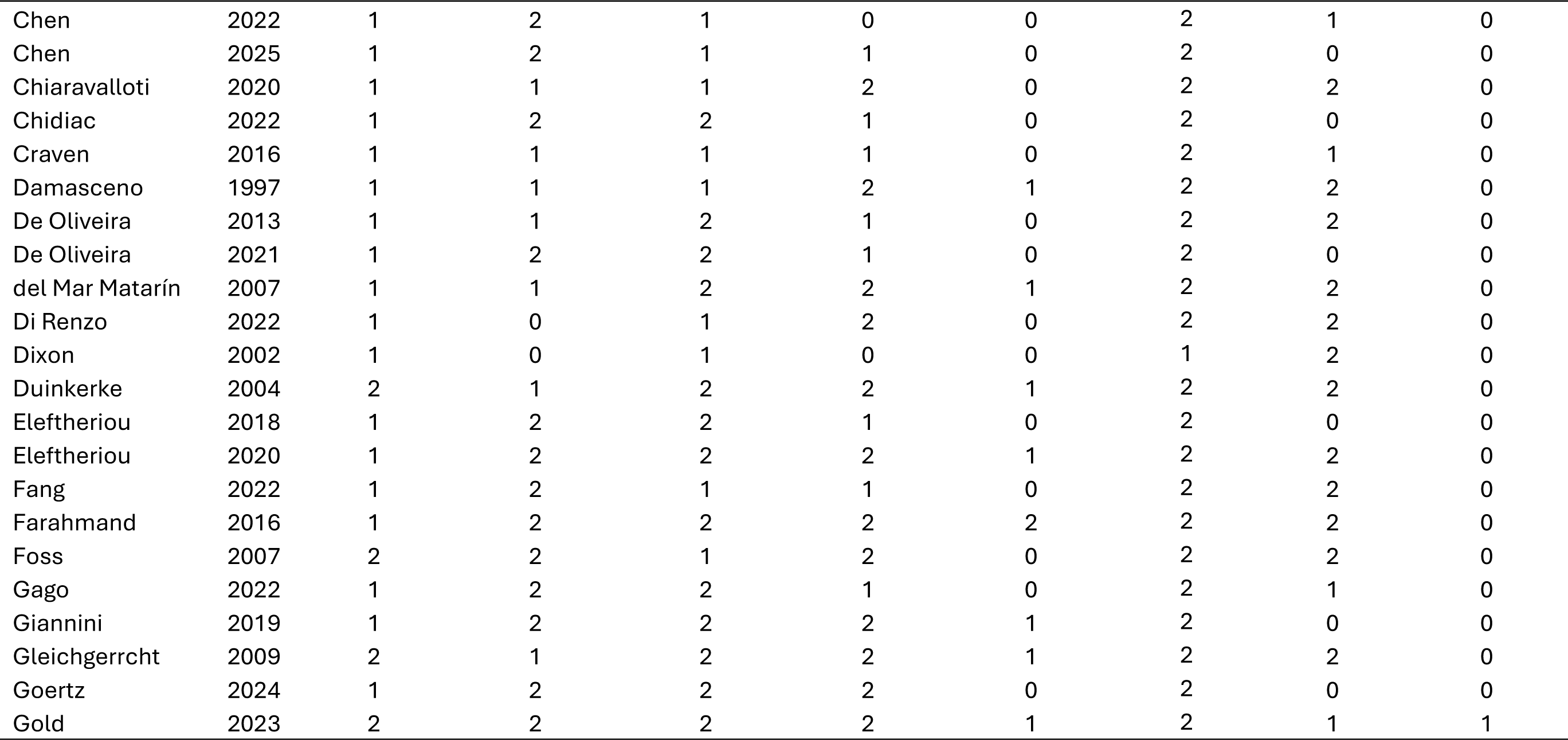
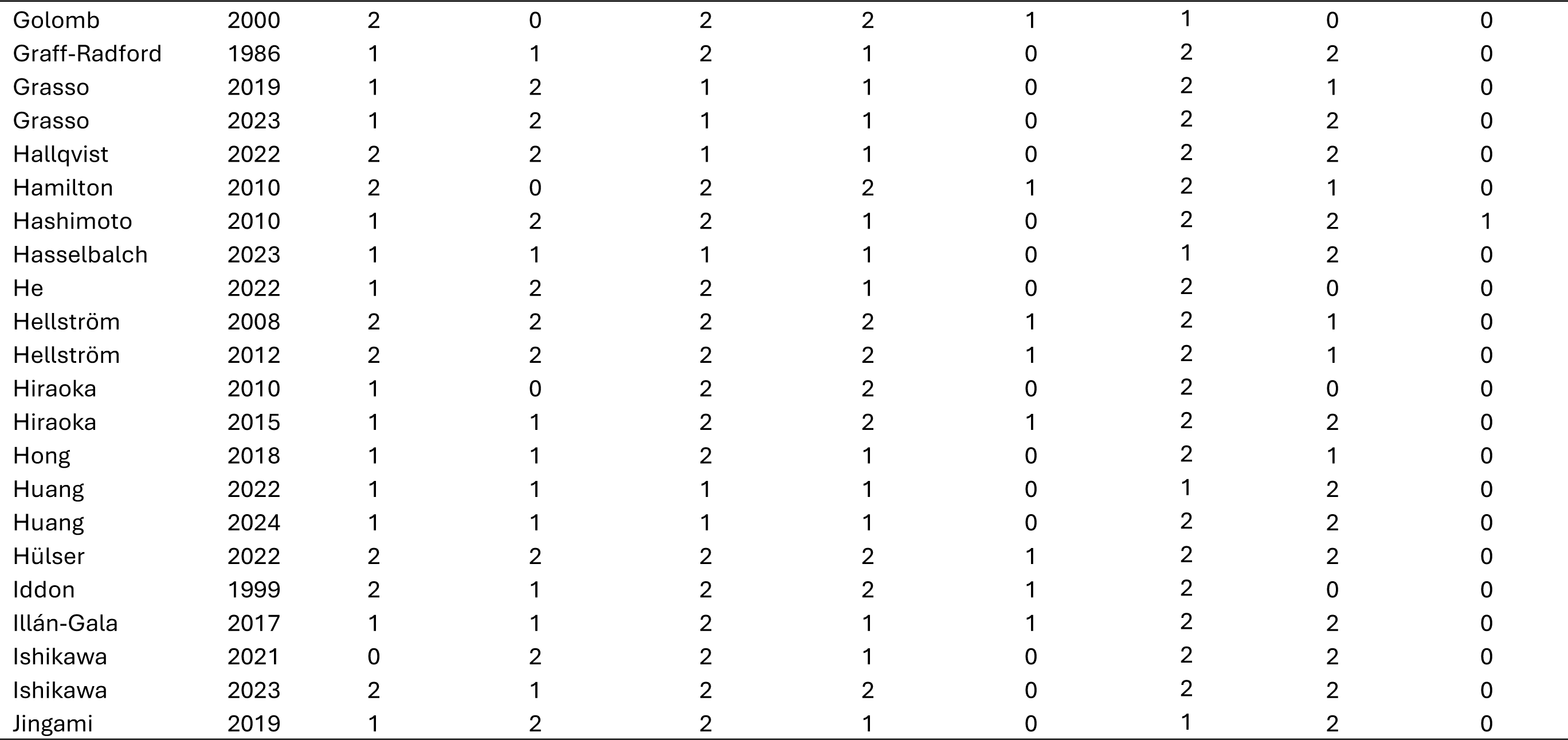
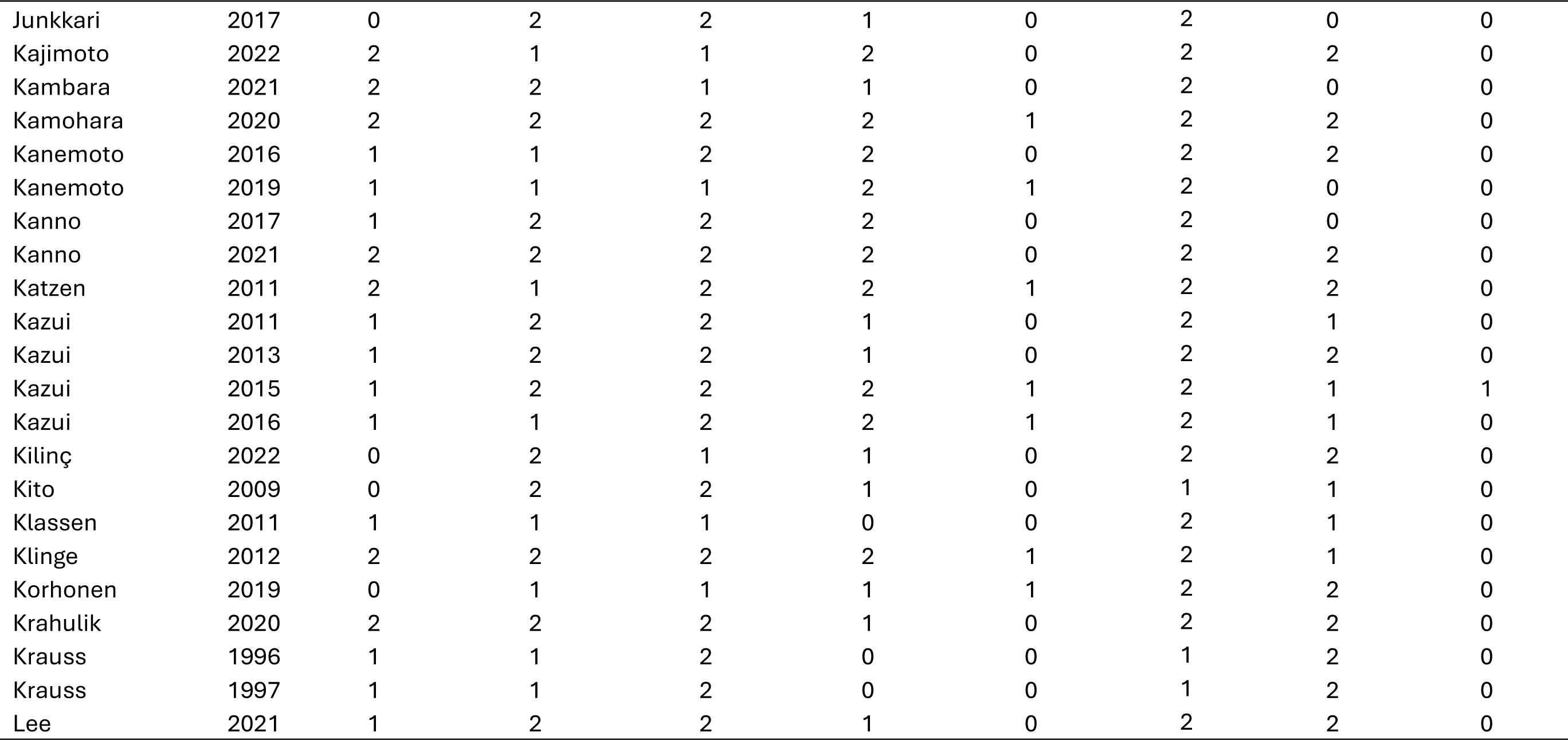
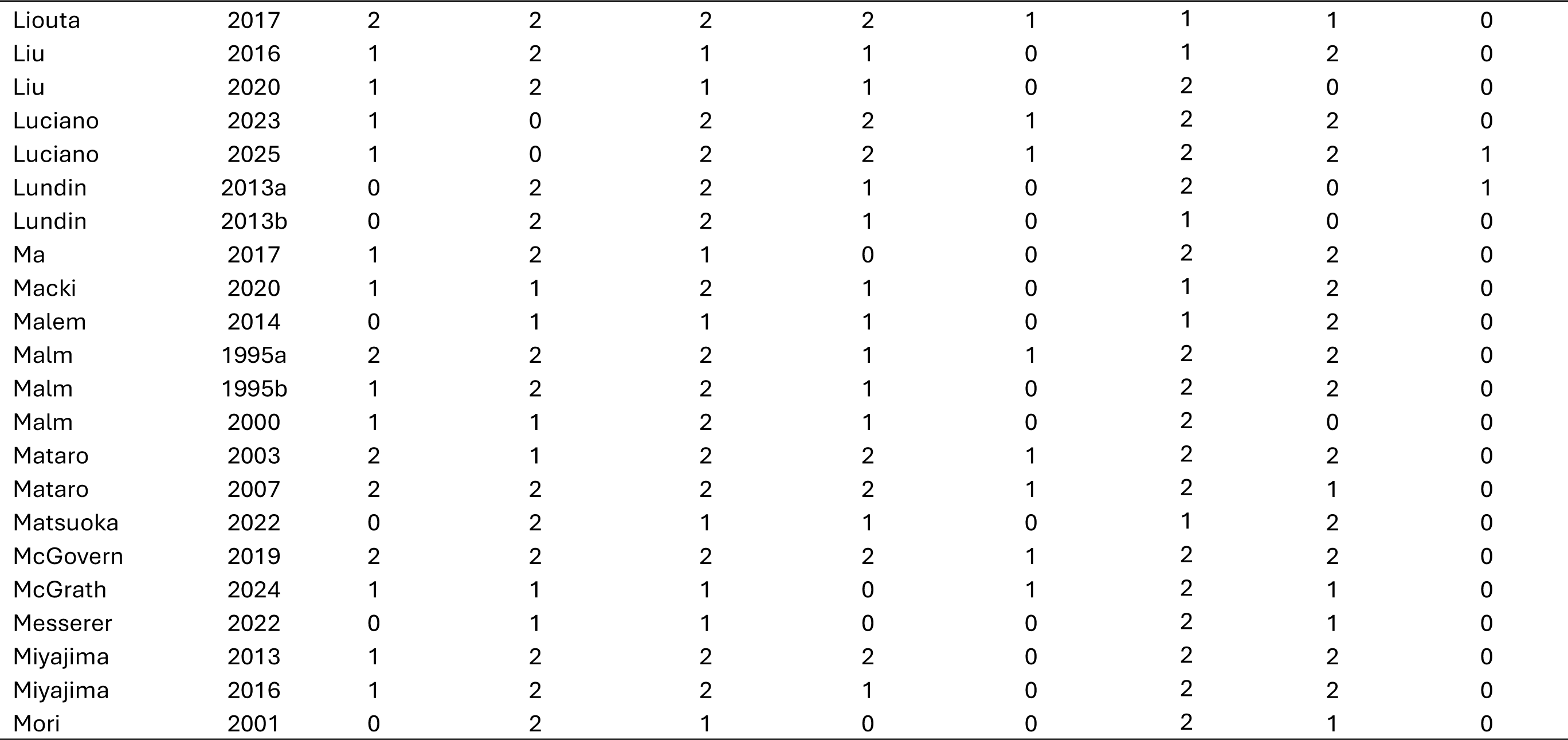
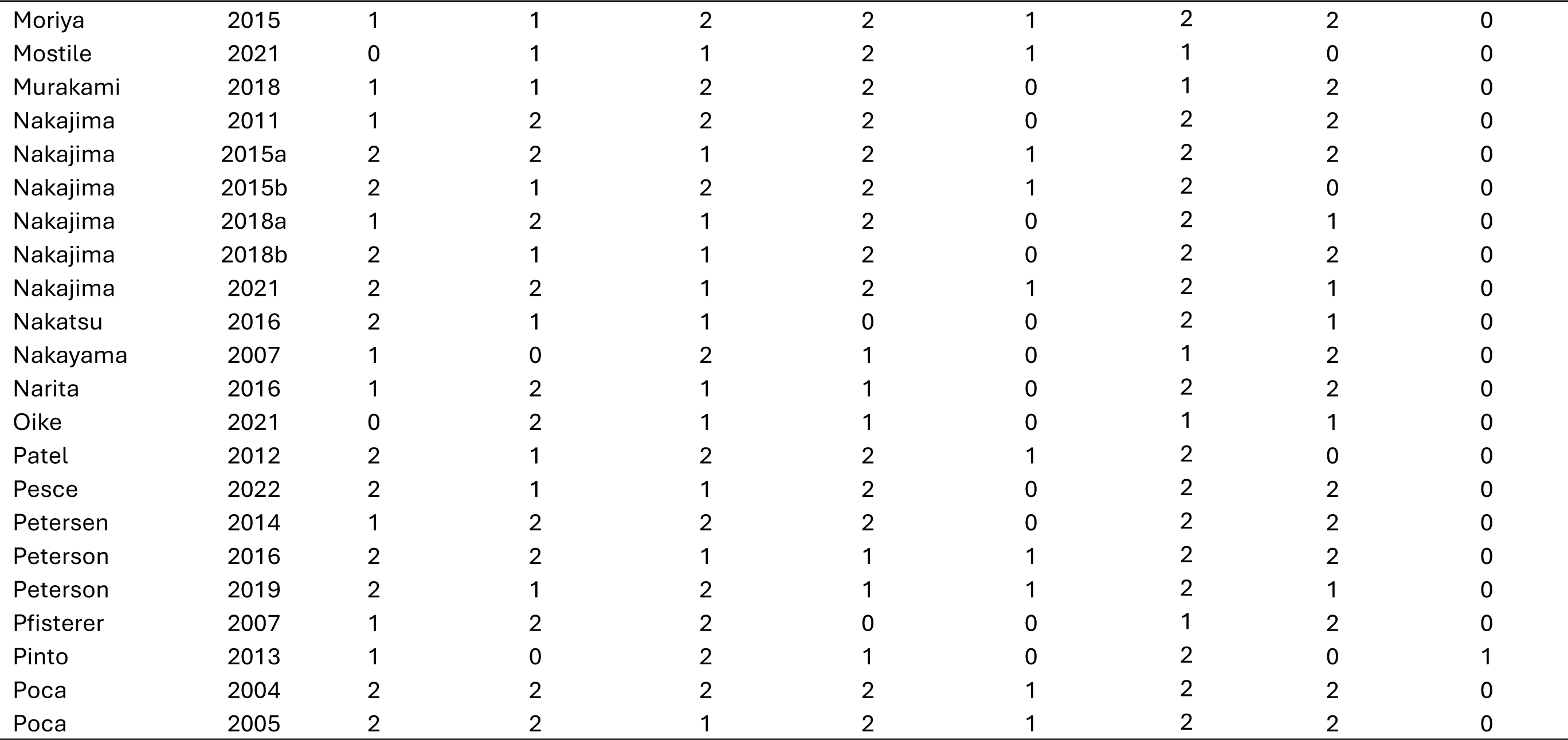
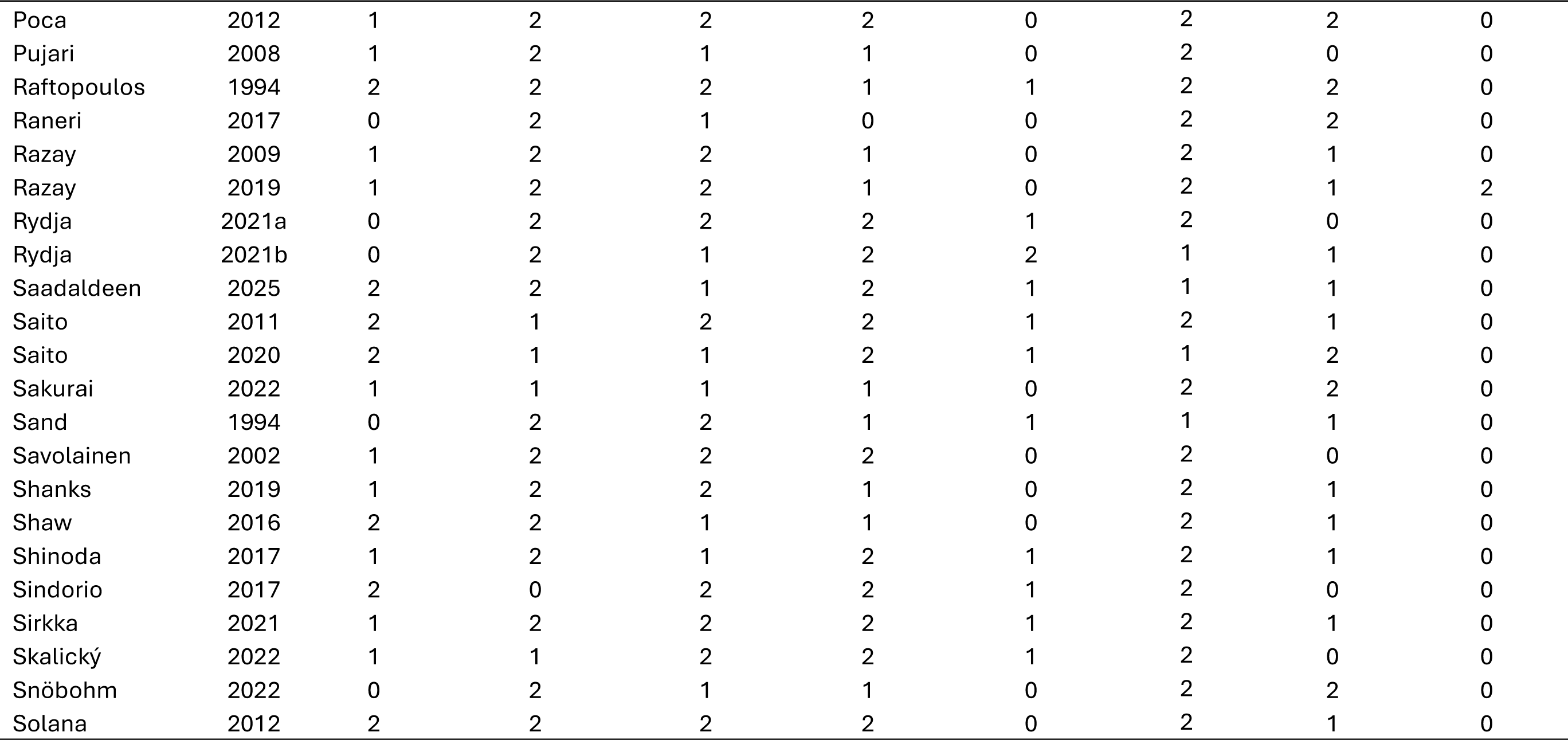
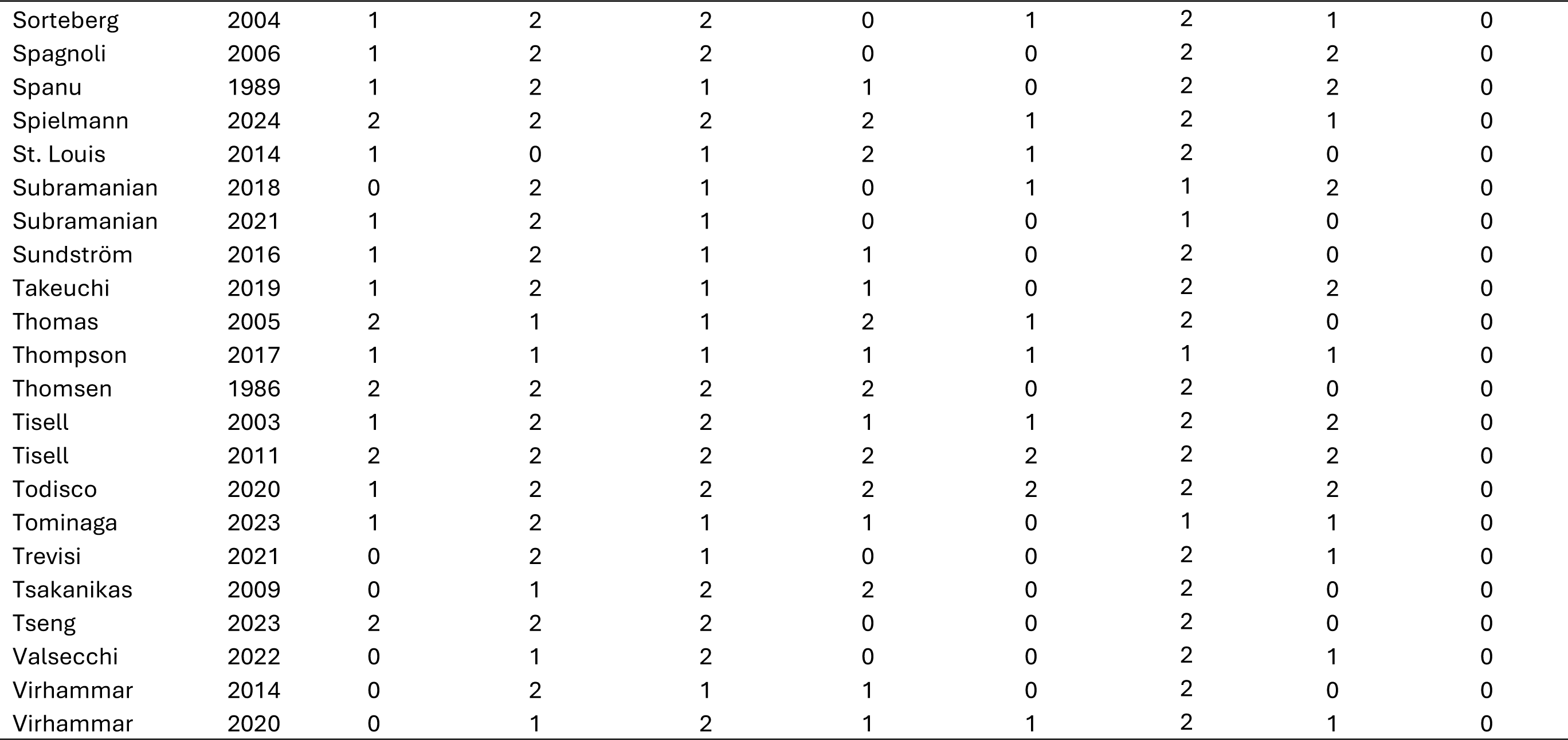
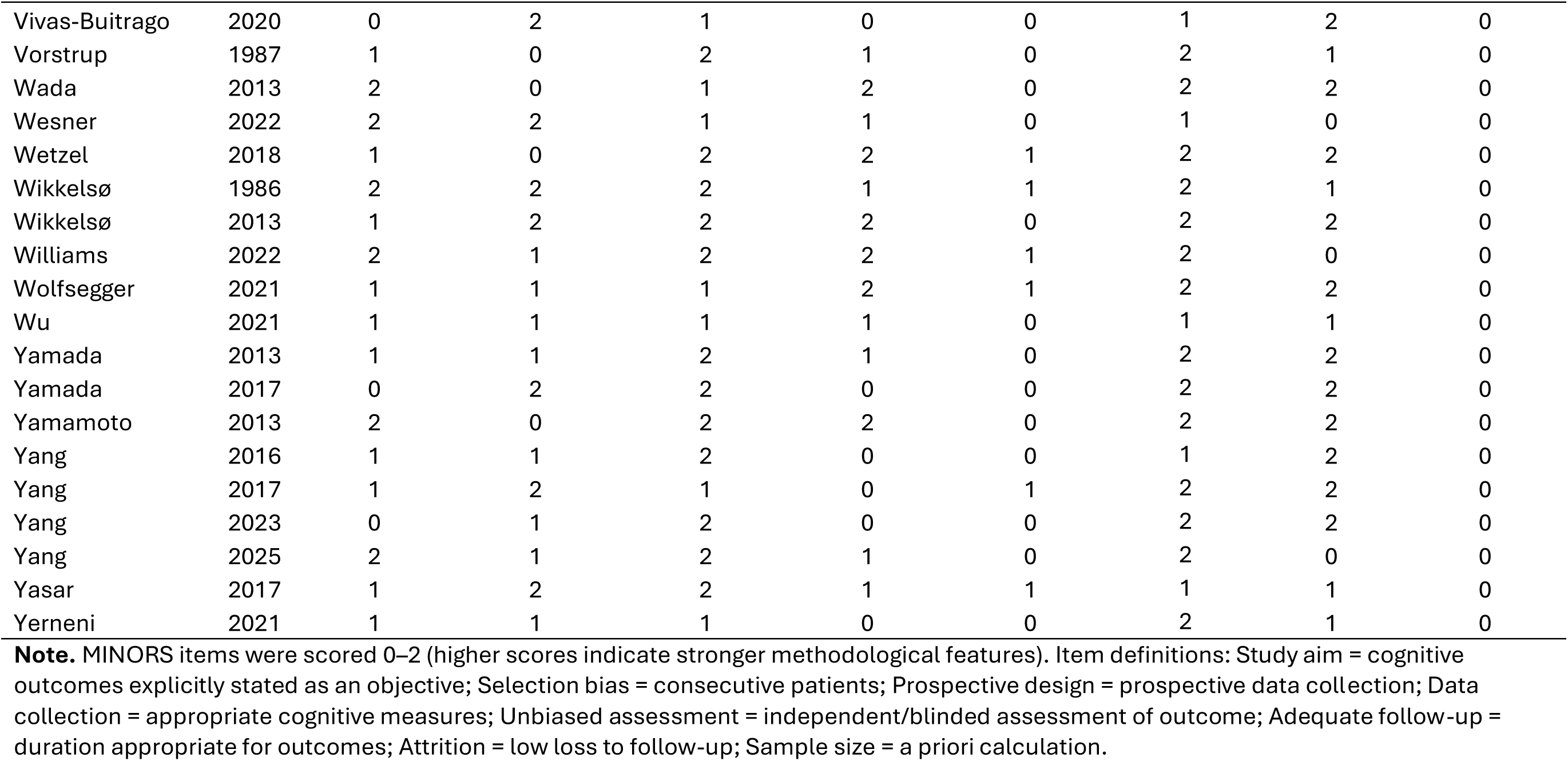
Domain-level MINORS quality assessment ratings for the 195 included studies.

**Supplementary Table 2.** Full list of cognitive tests reported in the 195 included studies, with reported cognitive outcome data.

| **Cognitive Test** | **Cognitive Domain / Test Type** | **Battery** | **Studies (n)** |
| --- | --- | --- | --- |
| Alzheimer's Disease Assessment Scale (ADAS) Comprehension of Spoken Language | Language | ADAS | 1 |
| ADAS Constructional Praxis | Visuospatial function | ADAS | 2 |
| ADAS Following Commands | Language | ADAS | 1 |
| ADAS Ideational Praxis | Executive function / Praxis | ADAS | 1 |
| ADAS Naming Objects and Fingers | Language | ADAS | 1 |
| ADAS Orientation | Orientation | ADAS | 1 |
| ADAS Remembering Test Instructions | Memory | ADAS | 1 |
| ADAS Spoken Language Ability | Language | ADAS | 1 |
| ADAS Word Recall | Memory | ADAS | 2 |
| ADAS Word Recognition | Memory | ADAS | 2 |
| ADAS Word-Finding Difficulty | Language | ADAS | 1 |
| Addenbrooke's Cognitive Exam (ACE) | Dementia screen |  | 2 |
| Alphabet Writing (Timed, Non-standardised Test) | Psychomotor speed |  | 3 |
| Attention (Unspecified Test) | Attention |  | 1 |
| Barrage Test | Attention |  | 1 |
| Benton Face Recognition | Face recognition / Visual perception |  | 1 |
| Benton Judgement of Line Orientation | Visuospatial function |  | 1 |
| Benton Visual Retention Test | Memory |  | 1 |
| Bingley's Memory Test | Memory |  | 5 |
| Brief Mental Deterioration Battery (BMDB) Word List | Memory | BMDB | 1 |

**Supplementary Table 2.** (Continued)
| **Cognitive Test** | **Cognitive Domain / Test Type** | **Battery** | **Studies (n)** |
| --- | --- | --- | --- |
| BMDB Analogies | Executive function | BMDB | 1 |
| BMDB Barrage Test | Attention | BMDB | 1 |
| BMDB Visual Memory - Immediate Recall | Memory | BMDB | 1 |
| Boston Diagnostic Aphasia Examination (BDAS) Fluency | Language |  | 1 |
| Boston Naming Test | Language |  | 7 |
| Buschke-Fuld 10-Word List - Learning and Recall | Memory |  | 1 |
| Cambridge Neuropsychological Test Automated Battery (CANTAB) Intra-Extra Dimensional Set Shift | Executive function | CANTAB | 1 |
| CANTAB Pattern Recognition | Memory | CANTAB | 1 |
| CANTAB Spatial Recognition | Memory | CANTAB | 1 |
| CANTAB Spatial Span | Working memory | CANTAB | 1 |
| Consortium to Establish a Registry for Alzheimer's Disease (CERAD) Boston Naming Test | Language | CERAD | 2 |
| CERAD Constructional Praxis | Visuospatial function | CERAD | 2 |
| CERAD Constructional Praxis - Delayed Recall | Memory | CERAD | 2 |
| CERAD Semantic Fluency | Language / Verbal fluency | CERAD | 2 |
| CERAD Word List (10-item) Learning, Recall and Recognition | Memory | CERAD | 2 |
| Clock Copy Test | Visuospatial function |  | 1 |
| Clock Drawing Test | Visuospatial function |  | 4 |
| Computerized General Neuropsychological iNPH Test (CoGNIT) Choice Reaction Time | Reaction time | CoGNIT | 1 |
| CoGNIT Finger Tapping | Motor skill | CoGNIT | 1 |

**Supplementary Table 2.** (Continued)
| **Cognitive Test** | **Cognitive Domain / Test Type** | **Battery** | **Studies (n)** |
| --- | --- | --- | --- |
| CoGNIT Stroop Congruent | Processing speed | CoGNIT | 1 |
| CoGNIT Stroop Incongruent | Executive function | CoGNIT | 1 |
| CoGNIT Trails A | Processing speed / Psychomotor speed | CoGNIT | 1 |
| CoGNIT Trails B | Executive function | CoGNIT | 1 |
| CoGNIT Word List Learning, Delayed Recall and Recognition | Memory | CoGNIT | 1 |
| Copy Drawing with Landmarks Test | Visuospatial function |  | 1 |
| Corsi Block Tapping Test | Working memory |  | 1 |
| Cube Drawing | Visuospatial function |  | 1 |
| Cylinders Test (timed) | Executive function / Psychomotor speed |  | 1 |
| Delis-Kaplan Executive Function System (DKEFS) Colour Word Interference Test | Executive function | DKEFS | 1 |
| Dementia Rating Scale (DRS) Attention | Attention | DRS | 2 |
| DRS Conceptualisation | Executive function | DRS | 2 |
| DRS Construction | Visuospatial function | DRS | 2 |
| DRS Initiation / Perseveration | Executive function | DRS | 2 |
| DRS Memory | Memory | DRS | 2 |
| DemTect | Dementia screen |  | 2 |
| Design Copy (Unspecified Test) | Visuospatial function |  | 1 |
| Episodic Memory (Unspecified Test) | Memory |  | 1 |
| Facial Affect Test | Social cognition |  | 1 |
| Figure Copy (Unspecified Test) | Visuospatial function |  | 1 |

**Supplementary Table 2.** (Continued)
| **Cognitive Test** | **Cognitive Domain / Test Type** | **Battery** | **Studies (n)** |
| --- | --- | --- | --- |
| Finger Tapping Test | Motor speed |  | 6 |
| Frontal Assessment Battery (FAB) | Executive function screen |  | 35 |
| Fuld Object Memory Test | Memory |  | 1 |
| Goldstein and Scheerer Block Design | Executive function / Visuospatial function |  | 1 |
| Grooved Pegboard Test | Psychomotor speed |  | 27 |
| Guild Memory Test - Paragraph Recall | Memory |  | 1 |
| Hasegawa Dementia Rating Scale - Revised | Dementia screen |  | 1 |
| Hopkins Verbal Learning Test - Learning and Delayed Recall | Memory |  | 2 |
| Identical Forms Test | Visuospatial function |  | 5 |
| Intersecting Pentagons Copy | Visuospatial function |  | 1 |
| Judgement of Line Orientation | Visuospatial function |  | 2 |
| Kendrick Object Learning Test (KOLT) | Memory |  | 1 |
| Kohs Test | Visuospatial function |  | 1 |
| Line Tracing (Timed, Non-standardised Test) | Psychomotor speed |  | 7 |
| Luria Hand Sequencing | Executive function / Motor sequencing |  | 1 |
| Mefferd & Moran Perceptual Speed Test | Perceptual speed |  | 1 |
| Memory Recall (Unspecified Test) | Memory |  | 1 |
| Mental Deterioration Battery - Figure Copy | Visuospatial function |  | 1 |
| Mini-Mental State Exam (MMSE) | Dementia screen |  | 128 |
| Montreal Cognitive Assessment (MoCA) | Dementia screen |  | 9 |

**Supplementary Table 2.** (Continued)
| **Cognitive Test** | **Cognitive Domain / Test Type** | **Battery** | **Studies (n)** |
| --- | --- | --- | --- |
| Multiple Features Targets Cancellation | Attention |  | 1 |
| Neuropsychological Assessment Battery (NAB) Auditory Comprehension | Language | NAB | 1 |
| NAB Bill Payment | Language | NAB | 1 |
| NAB Categories | Executive function | NAB | 1 |
| NAB Daily Living Memory - Immediate Recall, Delayed Recall and Recognition | Memory | NAB | 1 |
| NAB Design Construction | Visuospatial function | NAB | 1 |
| NAB Digit Span Backward | Attention | NAB | 1 |
| NAB Digit Span Forward | Attention | NAB | 1 |
| NAB Dots | Attention | NAB | 1 |
| NAB Driving Scenes | Attention | NAB | 1 |
| NAB Figure Drawing Copy | Visuospatial function | NAB | 1 |
| NAB Figure Recall | Memory | NAB | 1 |
| NAB Judgement | Executive function | NAB | 1 |
| NAB List Learning - Immediate Recall, Delayed Recall, Recognition | Memory | NAB | 1 |
| NAB Map Reading | Visuospatial function | NAB | 1 |
| NAB Mazes | Executive function | NAB | 1 |
| NAB Naming | Language | NAB | 1 |
| NAB Number and Letters | Attention | NAB | 1 |
| NAB Oral Production | Language | NAB | 1 |
| NAB Orientation | Orientation | NAB | 1 |

**Supplementary Table 2.** (Continued)
| **Cognitive Test** | **Cognitive Domain / Test Type** | **Battery** | **Studies (n)** |
| --- | --- | --- | --- |
| NAB Reading Comprehension | Language | NAB | 1 |
| NAB Shape Learning - Immediate and Delayed Recognition | Memory | NAB | 1 |
| NAB Story Learning - Immediate and Delayed Recall | Memory | NAB | 1 |
| NAB Visual Discrimination | Visuospatial function | NAB | 1 |
| NAB Word Generation | Executive function | NAB | 1 |
| NAB Writing | Language | NAB | 1 |
| Neuropsychological Test Battery (Unspecified Tests) | Battery |  | 3 |
| NIH Dimensional Card Sort | Executive function | NIH Toolbox | 1 |
| NIH Flanker Inhibitory Control | Executive function | NIH Toolbox | 1 |
| Numbers Test (Unspecified Test) | Attention |  | 1 |
| Oktem Verbal Learning Processes - Learning, Recall and Recognition | Memory |  | 1 |
| Orientation test (Unspecified Test) | Orientation |  | 1 |
| Pentagon Copy (Timed, Non-standardised Test) | Psychomotor speed |  | 3 |
| Phonemic Fluency - FAS (English) | Executive function / Verbal fluency |  | 5 |
| Phonemic Fluency - FAS (Spanish / Catalan) | Executive function / Verbal fluency |  | 2 |
| Phonemic Fluency - FPL (Italian) | Executive function / Verbal fluency |  | 1 |
| Phonemic Fluency - Fu, A, Ni (Japanese) | Executive function / Verbal fluency |  | 1 |
| Phonemic Fluency - KAS (Turkish) | Executive function / Verbal fluency |  | 1 |
| Phonemic Fluency - NKP (Czech) | Executive function / Verbal fluency |  | 2 |
| Phonemic Fluency - P (Spanish) | Executive function / Verbal fluency |  | 1 |

**Supplementary Table 2.** (Continued)
| **Cognitive Test** | **Cognitive Domain / Test Type** | **Battery** | **Studies (n)** |
| --- | --- | --- | --- |
| Phonemic Fluency - PAS (Finnish) | Executive function / Verbal fluency |  | 1 |
| Phonemic Fluency - S (Spanish / Catalan) | Executive function / Verbal fluency |  | 1 |
| Phonemic Fluency - ΧΣΑ (Greek) | Executive function / Verbal fluency |  | 1 |
| Phonemic Fluency - Unspecified Letter(s) | Executive function / Verbal fluency |  | 4 |
| Phrase construction (Unspecified Test) | Language |  | 1 |
| Proverbs (Unspecified Test) | Executive function |  | 1 |
| Purdue Pegboard Test | Psychomotor speed |  | 4 |
| Raven's Coloured Progressive Matrices | Nonverbal IQ test |  | 3 |
| Reaction Time - Continuous | Reaction time |  | 1 |
| Reaction Time - Simple | Reaction time |  | 1 |
| Reaction Time - Target | Reaction time |  | 1 |
| Reaction Time (Unspecified Test) | Reaction time |  | 2 |
| Repeatable Battery for the Assessment of Neuropsychological Status (RBANS) Coding | Processing speed | RBANS | 1 |
| RBANS Digit Span | Attention | RBANS | 1 |
| RBANS Figure Copy | Visuospatial function | RBANS | 1 |
| RBANS Figure Recall | Memory | RBANS | 1 |
| RBANS Line Orientation | Visuospatial function | RBANS | 1 |
| RBANS List Learning | Memory | RBANS | 1 |
| RBANS Naming | Language | RBANS | 1 |
| RBANS Semantic Fluency | Language / Verbal fluency | RBANS | 1 |

**Supplementary Table 2.** (Continued)
| **Cognitive Test** | **Cognitive Domain / Test Type** | **Battery** | **Studies (n)** |
| --- | --- | --- | --- |
| RBANS Story Memory | Memory | RBANS | 1 |
| Rey Auditory Verbal Learning Test (RAVLT) - Learning, Recall and Recognition | Memory |  | 32 |
| Rey Complex Figure - Copy | Visuospatial function |  | 11 |
| Rey Complex Figure - 3-minute Recall and Delayed Recall | Memory |  | 11 |
| Rivermead Behavioural Memory Test (RBMT) Story Immediate and Delayed Recall | Memory |  | 3 |
| RBMT Picture Recognition | Memory |  | 1 |
| Semantic Fluency - Animals (Czech) | Language / Verbal fluency |  | 1 |
| Semantic Fluency - Animals (English) | Language / Verbal fluency |  | 1 |
| Semantic Fluency - Animals (Japanese) | Language / Verbal fluency |  | 1 |
| Semantic Fluency - Animals (Spanish / Catalan) | Language / Verbal fluency |  | 5 |
| Semantic Fluency - Animals (Spanish) | Language / Verbal fluency |  | 1 |
| Semantic Fluency - Animals (Turkish) | Language / Verbal fluency |  | 1 |
| Semantic Fluency - Vegetables | Language / Verbal fluency |  | 1 |
| Semantic Fluency - Unspecified Category | Language / Verbal fluency |  | 8 |
| Sentence Repetition | Language / Working memory |  | 1 |
| Serial Dotting Test | Psychomotor speed |  | 3 |
| Spatial Span (Unspecified Test) | Working memory |  | 1 |
| Stroop Colour Naming | Processing speed |  | 21 |
| Stroop Colour Word (Interference) Test | Executive function |  | 30 |
| Symbol Digit Modalities Test (SDMT) | Processing speed |  | 5 |

**Supplementary Table 2.** (Continued)
| **Cognitive Test** | **Cognitive Domain / Test Type** | **Battery** | **Studies (n)** |
| --- | --- | --- | --- |
| Tactile Verbal Memory (Unspecified Test) | Memory |  | 1 |
| Token Test | Language |  | 2 |
| Tracks Task | Psychomotor speed |  | 2 |
| Trail Making Test A (TMT-A) | Processing speed / Psychomotor speed |  | 37 |
| Trail Making Test B (TMT-B) | Executive function |  | 24 |
| Verbal Analogies Test | Executive function |  | 1 |
| Visual Discrimination Test - Complex Form | Visuospatial function |  | 1 |
| Visual Discrimination Test - Directional Discrimination | Visuospatial function |  | 1 |
| Visual Discrimination Test - Length and Size | Visuospatial function |  | 1 |
| Visual Discrimination Test - Overlapping Figures | Visuospatial function |  | 1 |
| Visual Discrimination Test - Visual Counting | Visuospatial function |  | 1 |
| Visual Gestalts - Learning and Retention | Memory |  | 1 |
| Visual Memory - Recall (Unspecified Test) | Memory |  | 2 |
| Visual Memory - Recognition (Unspecified Test) | Memory |  | 1 |
| Wechsler Adult Intelligence Scale (WAIS) Full Scale IQ | IQ test | WAIS | 3 |
| WAIS Block Design | Executive function / Visuospatial function | WAIS | 6 |
| WAIS Coding / Digit Symbol | Processing speed | WAIS | 6 |
| WAIS Comprehension | Language | WAIS | 1 |
| WAIS Digit Span (Total) | Attention | WAIS | 1 |
| WAIS Digit Span Backward | Attention / Working memory | WAIS | 12 |

**Supplementary Table 2.** (Continued)
| **Cognitive Test** | **Cognitive Domain / Test Type** | **Battery** | **Studies (n)** |
| --- | --- | --- | --- |
| WAIS Digit Span Forward | Attention | WAIS | 11 |
| WAIS Similarities | Language / Executive function | WAIS | 1 |
| WAIS Symbol Search | Processing speed | WAIS | 3 |
| WAIS Vocabulary | Language | WAIS | 1 |
| Wechsler Memory Scale (WMS) Designs - Immediate and Delayed Recall | Memory | WMS | 1 |
| WMS Digit Span Backward | Attention / Working memory | WMS | 13 |
| WMS Digit Span Forward | Attention | WMS | 13 |
| WMS Information and Orientation | Orientation | WMS | 5 |
| WMS Logical Memory - Immediate and Delayed Recall | Memory | WMS | 10 |
| WMS Mental Control | Attention / Working memory | WMS | 8 |
| WMS Paired Associates - Learning and Recall | Memory | WMS | 5 |
| WMS Visual Reproduction - Immediate and Delayed Recall | Memory | WMS | 8 |
| Western Aphasia Battery Object Naming | Language |  | 1 |
| Word List Immediate Recall, Delayed Recall and Recognition (Unspecified Test) | Memory |  | 4 |

**Note.** Tests with multiple editions or alternate forms have been grouped under a unified heading, where applicable (e.g. WAIS, WMS); Unspecified Test = study did not provide the exact test name or version; The listed cognitive domain reflects either the primary domain classified in the source article or the most commonly accepted classification.

**Supplementary Table 3.**
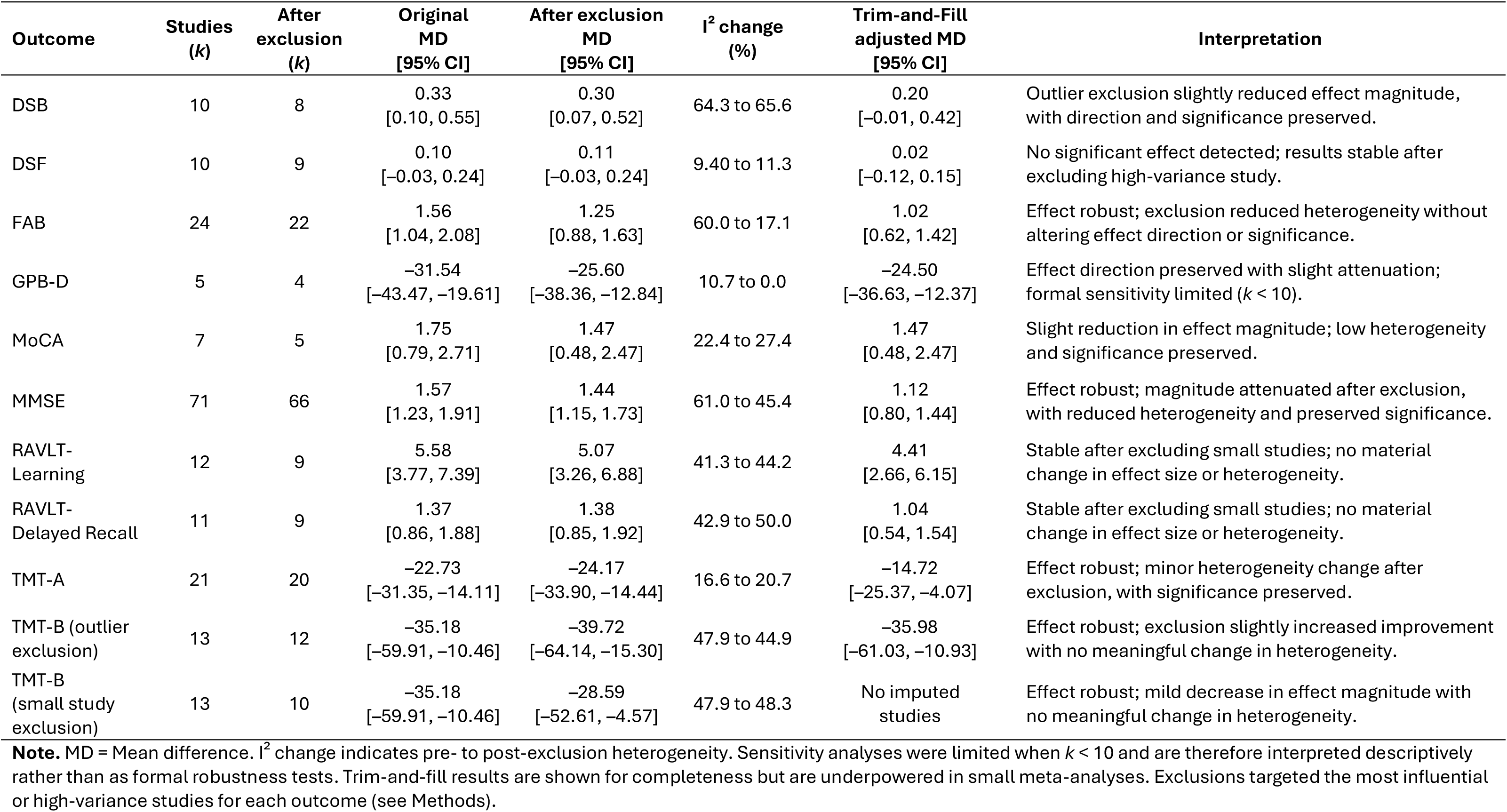
Summary of sensitivity analyses for cognitive outcomes following shunt surgery in iNPH.

| Outcome | Studies<br>(*k*) | After<br>exclusion<br>(*k*) | Original<br>MD<br>[95% CI] | After exclusion<br>MD<br>[95% CI] | I<sup>2</sup> change<br>(%) | Trim-and-Fill<br>adjusted MD<br>[95% CI] | Interpretation |
| --- | --- | --- | --- | --- | --- | --- | --- |
| DSB | 10 | 8 | 0.33<br>[0.10, 0.55] | 0.30<br>[0.07, 0.52] | 64.3 to 65.6 | 0.20<br>[−0.01, 0.42] | Outlier exclusion slightly reduced effect magnitude, with direction and significance preserved. |
| DSF | 10 | 9 | 0.10<br>[−0.03, 0.24] | 0.11<br>[−0.03, 0.24] | 9.40 to 11.3 | 0.02<br>[−0.12, 0.15] | No significant effect detected; results stable after excluding high-variance study. |
| FAB | 24 | 22 | 1.56<br>[1.04, 2.08] | 1.25<br>[0.88, 1.63] | 60.0 to 17.1 | 1.02<br>[0.62, 1.42] | Effect robust; exclusion reduced heterogeneity without altering effect direction or significance. |
| GPB-D | 5 | 4 | −31.54<br>[−43.47, −19.61] | −25.60<br>[−38.36, −12.84] | 10.7 to 0.0 | −24.50<br>[−36.63, −12.37] | Effect direction preserved with slight attenuation; formal sensitivity limited (*k* < 10). |
| MoCA | 7 | 5 | 1.75<br>[0.79, 2.71] | 1.47<br>[0.48, 2.47] | 22.4 to 27.4 | 1.47<br>[0.48, 2.47] | Slight reduction in effect magnitude; low heterogeneity and significance preserved. |
| MMSE | 71 | 66 | 1.57<br>[1.23, 1.91] | 1.44<br>[1.15, 1.73] | 61.0 to 45.4 | 1.12<br>[0.80, 1.44] | Effect robust; magnitude attenuated after exclusion, with reduced heterogeneity and preserved significance. |
| RAVLT-Learning | 12 | 9 | 5.58<br>[3.77, 7.39] | 5.07<br>[3.26, 6.88] | 41.3 to 44.2 | 4.41<br>[2.66, 6.15] | Stable after excluding small studies; no material change in effect size or heterogeneity. |
| RAVLT-Delayed Recall | 11 | 9 | 1.37<br>[0.86, 1.88] | 1.38<br>[0.85, 1.92] | 42.9 to 50.0 | 1.04<br>[0.54, 1.54] | Stable after excluding small studies; no material change in effect size or heterogeneity. |
| TMT-A | 21 | 20 | −22.73<br>[−31.35, −14.11] | −24.17<br>[−33.90, −14.44] | 16.6 to 20.7 | −14.72<br>[−25.37, −4.07] | Effect robust; minor heterogeneity change after exclusion, with significance preserved. |
| TMT-B (outlier exclusion) | 13 | 12 | −35.18<br>[−59.91, −10.46] | −39.72<br>[−64.14, −15.30] | 47.9 to 44.9 | −35.98<br>[−61.03, −10.93] | Effect robust; exclusion slightly increased improvement with no meaningful change in heterogeneity. |
| TMT-B (small study exclusion) | 13 | 10 | −35.18<br>[−59.91, −10.46] | −28.59<br>[−52.61, −4.57] | 47.9 to 48.3 | No imputed studies | Effect robust; mild decrease in effect magnitude with no meaningful change in heterogeneity. |

**Note.** MD = Mean difference. I<sup>2</sup> change indicates pre- to post-exclusion heterogeneity. Sensitivity analyses were limited when *k* < 10 and are therefore interpreted descriptively rather than as formal robustness tests. Trim-and-fill results are shown for completeness but are underpowered in small meta-analyses. Exclusions targeted the most influential or high-variance studies for each outcome (see Methods).

## Supplementary Text S3. Sensitivity analyses interpretive summary

Sensitivity analyses summarised in Supplementary Table 3 confirmed the robustness of pooled cognitive effects across all outcomes. Stepwise exclusion of small, high-variance, or directionally inconsistent studies did not materially alter the direction or significance of any pooled estimates. For key outcomes, TMT-A and TMT-B remained significant after exclusion of influential datasets, indicating consistent improvements in processing speed/psychomotor speed and executive function. In TMT-B, removal of an outlier dataset (Poca et al., 2004) increased the magnitude of the pooled effect (−35.18s to −39.72s) without altering heterogeneity. For GPB-D, exclusion of the largest study (Hellstrom et al., 2012) modestly reduced the pooled mean difference (–31.5s to –25.6s) but did not alter its direction or significance, supporting robustness despite the small number of studies. Across all remaining measures (DSB, DSF, FAB, MoCA, MMSE, RAVLT Learning, RAVLT Delayed Recall), exclusion of high-influence or outlier studies minimally affected pooled estimates and slightly reduced heterogeneity. Trim-and-fill analyses indicated limited funnel-plot asymmetry and minor attenuation of pooled effects, supporting the overall stability of the findings.

**Supplementary Figure 1.**
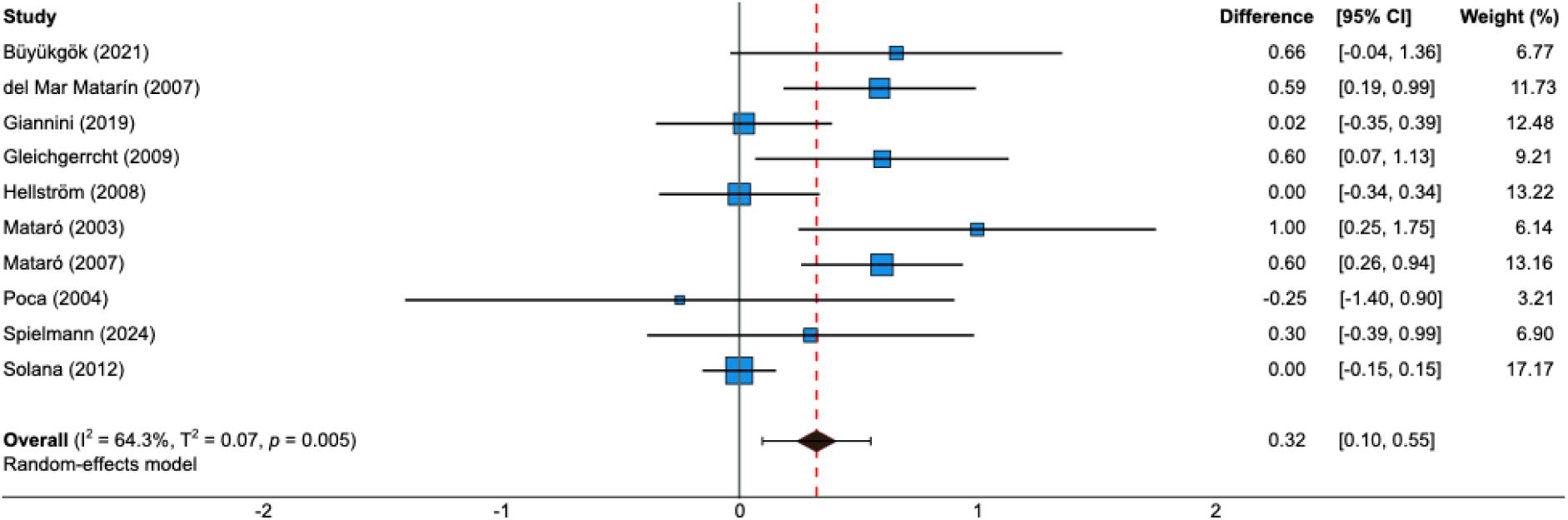
Digit Span Backward (DSB) **Note.** The forest plot illustrates the difference in DSB scores post-shunt surgery. The pooled mean difference was calculated using a random-effects model. Individual study estimates are presented with their 95% confidence intervals. Average differences are also shown with 95% confidence intervals. Unit of measurement = Maximum number of digits recalled in reverse order.

**Supplementary Figure 2.**
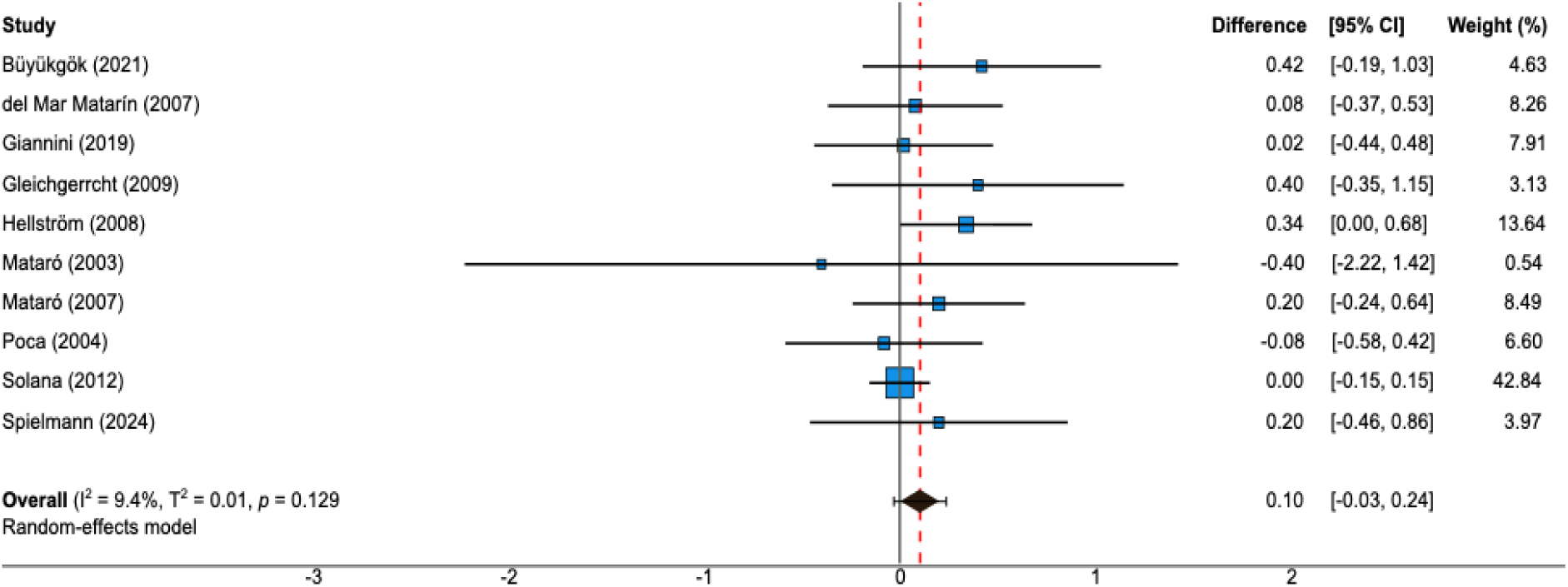
Digit Span Forward (DSF) **Note.** The forest plot illustrates the difference in DSF scores post-shunt surgery. The pooled mean difference was calculated using a random-effects model. Individual study estimates are presented with their 95% confidence intervals. Average differences are also shown with 95% confidence intervals. Unit of measurement = Maximum number of digits recalled.

**Supplementary Figure 3.**
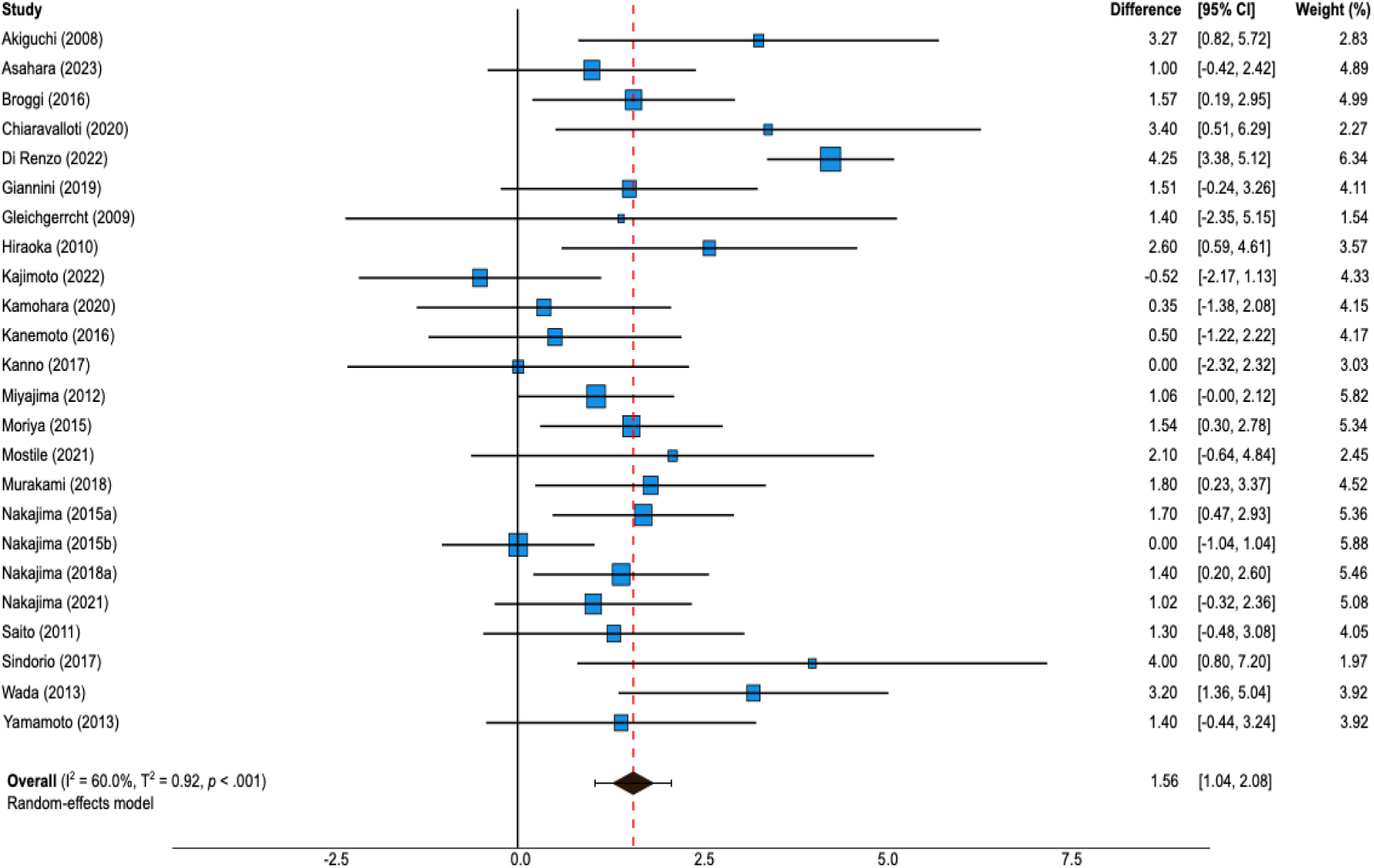
Frontal Assessment Battery (FAB) **Note.** The forest plot illustrates the difference in FAB scores post-shunt surgery. The pooled mean difference was calculated using a random-effects model. Individual study estimates are presented with their 95% confidence intervals. Average differences are also shown with 95% confidence intervals. Unit of measurement = Raw score based on a maximum of 18.

**Supplementary Figure 4.**
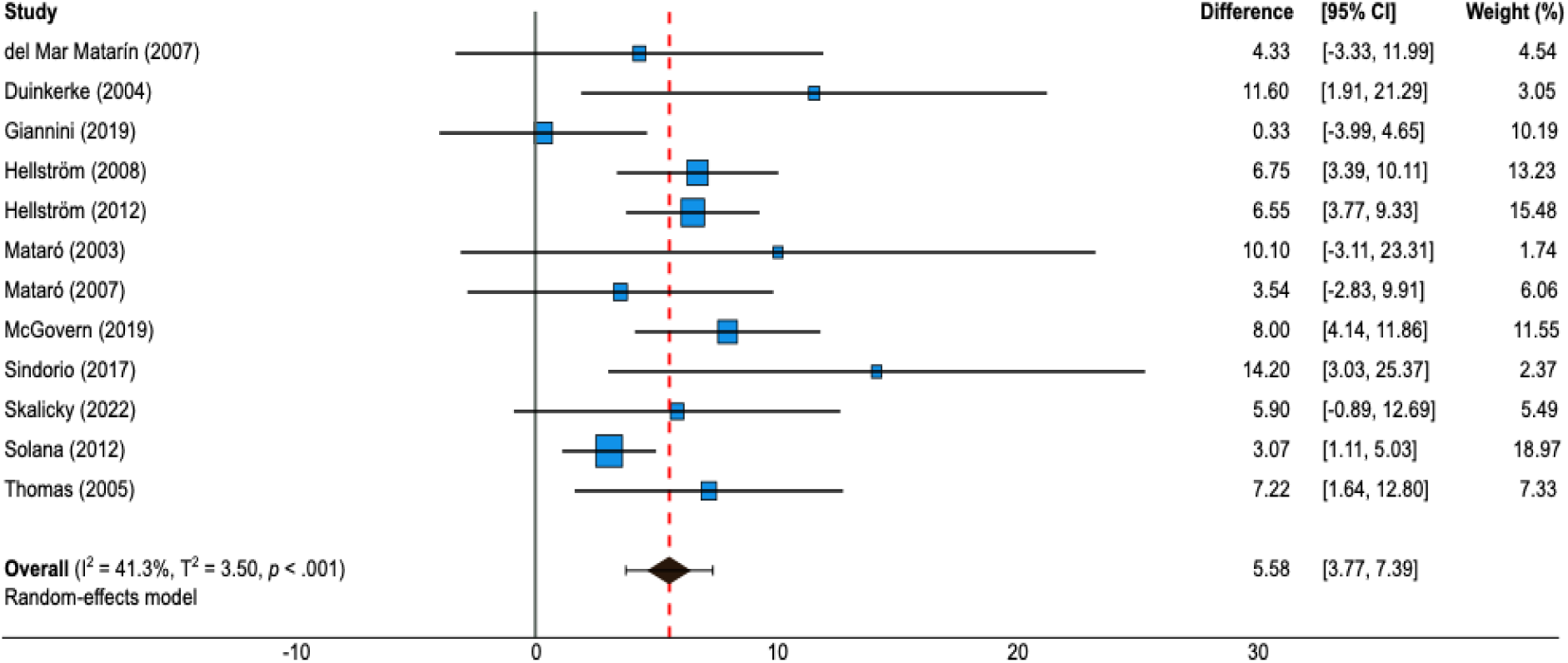
Rey Auditory Verbal Learning Test (RAVLT) Learning. **Note.** The forest plot illustrates the difference in RAVLT Learning Trials scores post-shunt surgery. The pooled mean difference was calculated using a random-effects model. Individual study estimates are presented with their 95% confidence intervals. Average differences are also shown with 95% confidence intervals. Unit of measurement = Total number of words recalled during the learning trials.

**Supplementary Figure 5.**
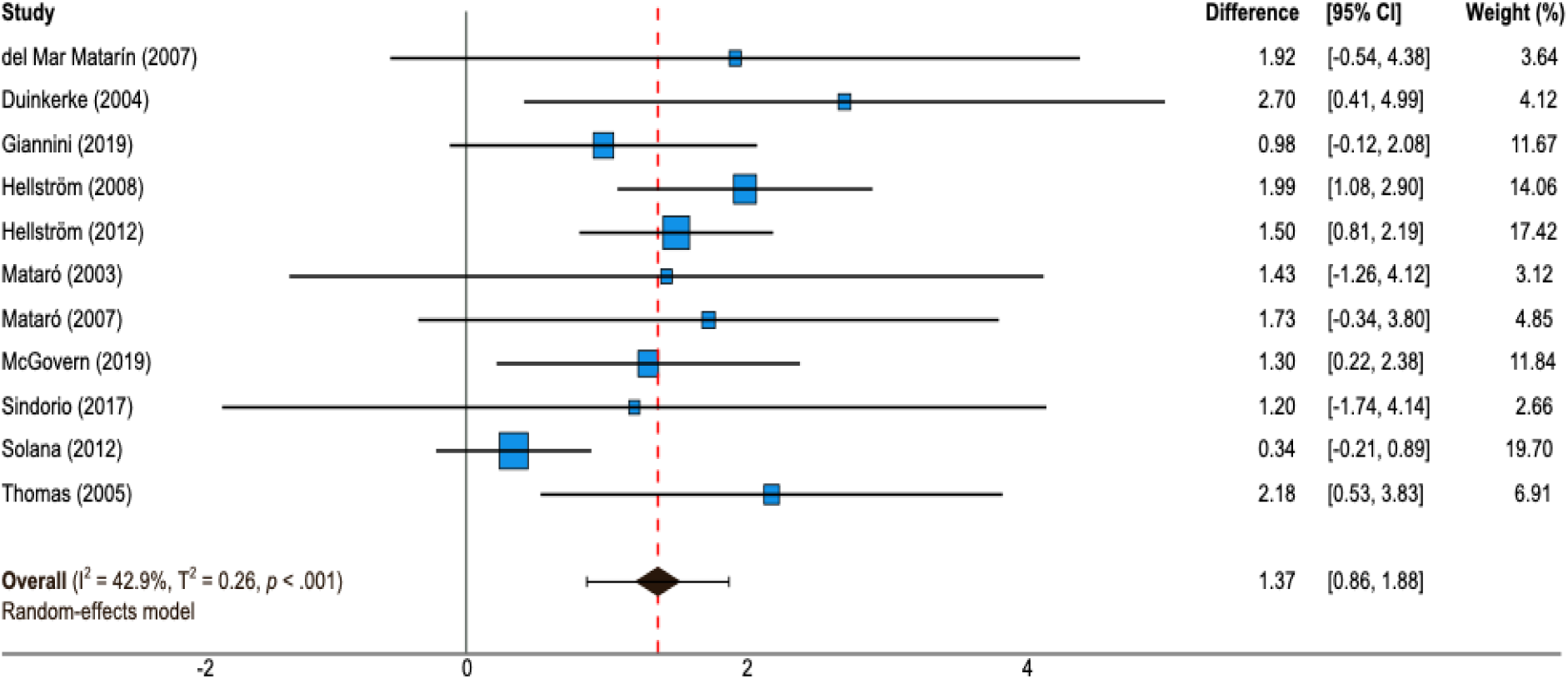
Rey Auditory Verbal Learning Test (RAVLT) Delayed Recall. **Note.** The forest plot illustrates the difference in RAVLT Delayed Recall scores post-shunt surgery. The pooled mean difference was calculated using a random-effects model. Individual study estimates are presented with their 95% confidence intervals. Average differences are also shown with 95% confidence intervals. Unit of measurement = Total number of words recalled following a delay, based on a maximum of 15.

**Supplementary Figure 6.**
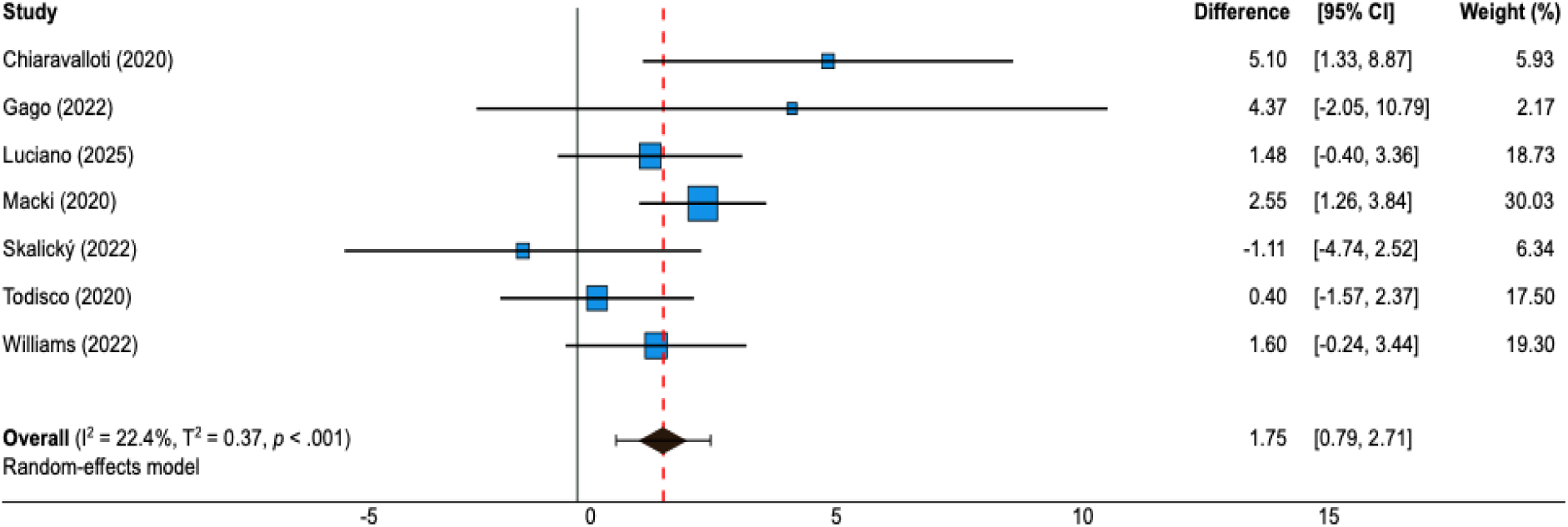
Montreal Cognitive Assessment (MoCA) **Note.** The forest plot illustrates the difference in MoCA scores post-shunt surgery. The pooled mean difference was calculated using a random-effects model. Individual study estimates are presented with their 95% confidence intervals. Average differences are also shown with 95% confidence intervals. Unit of measurement = Raw score based on a maximum of 30.

**Supplementary Figure 7.**
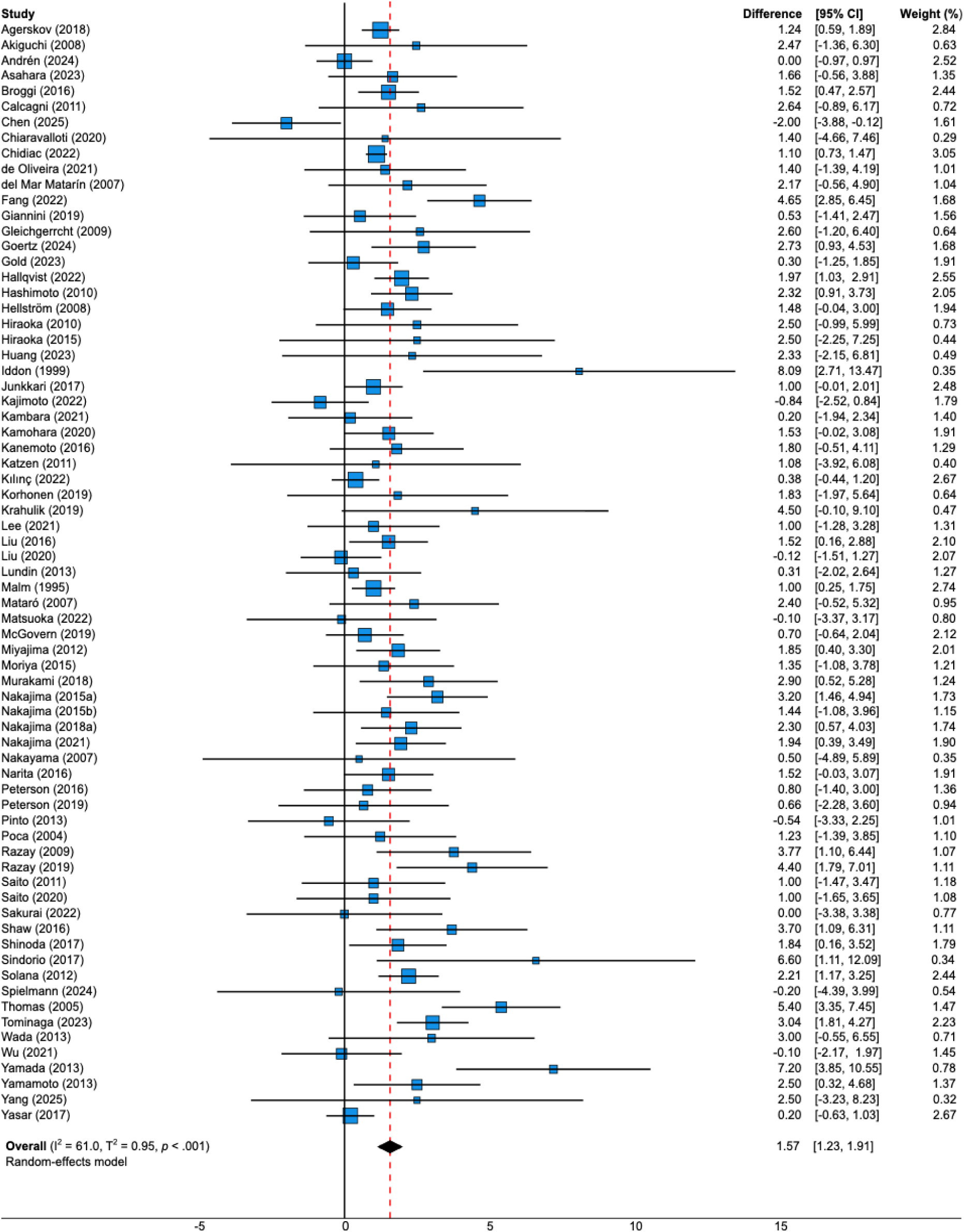
Mini-Mental State Exam (MMSE) **Note.** The forest plot illustrates the difference in MMSE scores post-shunt surgery. The pooled mean difference was calculated using a random-effects model. Individual study estimates are presented with their 95% confidence intervals. Average differences are also shown with 95% confidence intervals. Unit of measurement = Raw score based on a maximum of 30.

